# Blood Distribution of SARS-CoV-2 Lipid Nanoparticle mRNA Vaccine in Humans

**DOI:** 10.1101/2024.07.25.24311039

**Authors:** Stephen J. Kent, Shiyao Li, Thakshila H. Amarasena, Arnold Reynaldi, Wen Shi Lee, Michael G. Leeming, David H. O’Connor, Julie Nguyen, Helen E. Kent, Frank Caruso, Jennifer A. Juno, Adam K. Wheatley, Miles P. Davenport, Yi Ju

**Affiliations:** Department of Microbiology and Immunology, Peter Doherty Institute for Infection and Immunity, The University of Melbourne, Melbourne, Victoria 3000, Australia; Melbourne Sexual Health Centre and Department of Infectious Diseases, Alfred Hospital and Central Clinical School, Monash University, Melbourne, Victoria 3000, Australia; School of Science, RMIT University, Melbourne, Victoria 3000, Australia; Infection Analytics Program, Kirby Institute for Infection and Immunity, University of New South Wales, Sydney, New South Wales 2052, Australia; Melbourne Mass Spectrometry and Proteomics Facility, The Bio21 Molecular Science and Biotechnology Institute, The University of Melbourne, Parkville, Victoria 3010, Australia; Department of Chemical Engineering, The University of Melbourne, Melbourne, Victoria 3000, Australia

**Author notes:** These authors contributed equally (Stephen J. Kent, Shiyao Li).

**Keywords:** PEGylated lipid nanoparticle, COVID-19, kinetics of mRNA, immunoglobulins, biomolecular coronas, particle–immune cell interactions

## Abstract

Lipid nanoparticle mRNA vaccines are an exciting but new technology used in humans. There is limited understanding of factors that influence their biodistribution and immunogenicity. Antibodies to polyethylene glycol (PEG), which is on the surface of the lipid nanoparticle, are detectable in humans and boosted by human mRNA vaccination. We hypothesized that PEG-specific antibodies could increase the clearance of mRNA vaccines. We developed methods to quantify both the mRNA and ionizable lipid in frequent serial blood samples from 19 subjects receiving Moderna SPIKEVAX mRNA booster immunization. Both the mRNA and ionizable lipid peaked in blood 1-2 days post vaccination (median peak level 0.19 and 3.22 ng mL^-1^, respectively). The mRNA was detectable out to 14-28 days post-vaccination in most subjects. We measured the proportion of mRNA that was relatively intact in blood over time and found the decay kinetics of the intact mRNA and ionizable lipid were identical, suggesting the intact lipid nanoparticle recirculates in blood. However, mRNA and ionizable lipid decay rates did not correlate with baseline levels of PEG-specific nor spike-specific antibodies. The magnitude of mRNA and ionizable lipid detected in blood did correlate with the boost in PEG antibodies. Further, the ability of subject’s monocytes to phagocytose lipid nanoparticles had an inverse relationship with the rise in PEG antibodies. This suggests circulation of mRNA lipid nanoparticle vaccines into the blood and their ability to be cleared by phagocytes influence PEG immunogenicity of mRNA vaccines. Overall, this work defines the pharmacokinetics of lipid nanoparticle mRNA vaccine components in human blood after intramuscular injection and the factors that influence this. These insights should prove useful in improving the future safety and efficacy of lipid nanoparticle mRNA vaccines and therapeutics.

---

Lipid nanoparticle mRNA vaccines have revolutionized vaccinology and saved many lives during the COVID-19 pandemic.^1^ Lipid nanoparticle mRNA vaccines typically contain 5 materials – mRNA, an ionizable lipid, a polyethylene glycol (PEG)–lipid, a helper lipid and cholesterol. Although the vaccines are delivered intramuscularly (IM) and act primarily in the draining lymph node, recent studies have suggested that at least small amounts of the mRNA vaccines may distribute in humans more widely than originally anticipated. A primarily cross-sectional study detected mRNA in blood for up to 15 days after mRNA vaccination.^2^ Low levels of the vaccine mRNA were detected in breast milk up to 45 hours post-vaccination.^3,4^ An autopsy study of people dying incidentally after vaccination found mRNA in tissues (axillary lymph nodes and heart) up to 30 days after vaccination.^5^ Presumably the mRNA reached breast milk and tissues following circulation in blood. Despite evidence in animals and humans that mRNA can be detected in blood after vaccination,^2,6^ studies of the pharmacokinetics of mRNA lipid nanoparticle components in blood in humans are lacking.

Vaccines that contain non-human materials other than the vaccine antigens, such as adenovirus vectors, can induce immune responses to those products, known as anti-vector responses. If strong enough, such anti-vector responses can clear the vaccine more rapidly, limiting the immunogenicity of booster vaccinations or potentially causing other unintended effects.^7,8^ We recently found that mRNA vaccines can boost PEG-specific antibodies in humans,^9^ confirmed by multiple groups.^10–13^ PEG is however relatively weakly immunogenic and the PEG-specific antibodies remained relatively low (endpoint binding titer generally <10^3^) after 2 mRNA vaccinations and did not influence the immunogenicity of the vaccines.^9^ None-the-less, the long-term consequences of boosting PEG-specific antibodies after multiple mRNA vaccinations are unknown.^14^ In particular, even the modest boost in anti-PEG antibodies resulted in detectable increases in the ability of human blood monocytes to phagocytose nanomaterials containing PEG in vitro.^9^ This may be a greater concern for the clearance of PEGylated nanomaterials administered intravenously in contrast to IM-delivered mRNA vaccines which primarily act at the draining lymph node rather than circulating more widely.

We hypothesized that we could quantify the decay kinetics of small amounts of lipid nanoparticle mRNA vaccines that spill into the blood, and that the decay rates would be influenced by levels of PEG antibody levels. We further hypothesized that vaccine mRNA immunogenicity and the capacity of monocytes to phagocytose lipid nanoparticles might also be influenced by lipid nanoparticle levels or PEG antibodies in blood. To evaluate this, we studied IM-delivered mRNA vaccines in a cohort of 19 humans through serially sampling plasma early after vaccination and developing novel methods to quantify vaccine mRNA by PCR and the unique ionizable lipid by mass spectrometry (Figure 1a).

**Figure 1.**
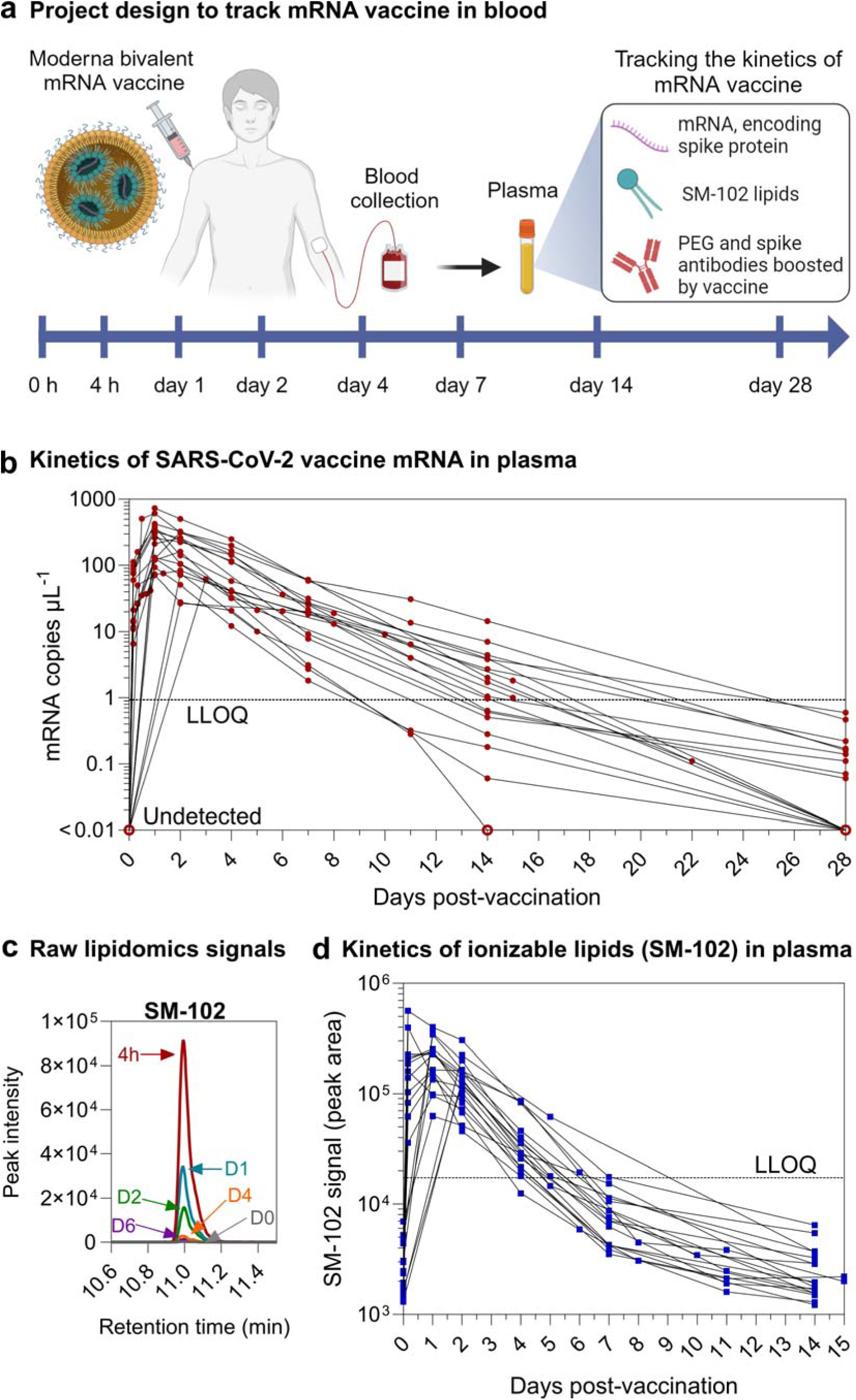
In vivo kinetics of mRNA and ionizable lipid from the SPIKEVAX SARS-CoV-2 mRNA vaccine in human blood. **a** Schematic illustration of the project design to track the kinetics of mRNA vaccine in human blood of 19 healthy subjects who received one dose of Moderna SPIKEVAX COVID-19 bivalent mRNA vaccine (see Table S1 for subject information). Plasma samples were collected at pre-vaccination (day 0) and at a median of 8 (range 4–14) other timepoints between 4 hours and 28 days post-vaccination. Created with BioRender.com. **b** Longitudinal mRNA levels in the plasma of the 19 subjects. The lower limit of quantification (LLOQ) is shown as a dashed line. The undetected samples were plotted at <0.01 copies µL^-1^ with open red circles. To improve readability, the detailed mRNA kinetics within the first 24 hours of vaccination is shown in Figure S2. **c** Representative image showing the peak intensity of SM-102 signals in a set of plasma samples from a subject at day 0–6 post-vaccination determined by the mass spectrometry-based lipidomics. **d** Longitudinal SM-102 ionizable lipid levels in the plasma of the 19 subjects. The LLOQ is shown as a dashed line.

## RESULTS AND DISCUSSION

### Human subjects

We studied 19 subjects receiving a bivalent Moderna SPIKEVAX booster immunization. The subjects ranged from 24–70 (mean 42) years old with 63% female and had received 3–4 (median 3) dose of monovalent COVID-19 vaccines before receiving the bivalent mRNA vaccine (details of subjects and vaccines in Table S1). Serial blood samples (on median 9, range 5–15 samples/subject) were collected pre-vaccination and from 4 hours to 28 days post-vaccination. Plasma (with EDTA or heparin anticoagulation) were stored within 4 hours at -80°C and PBMC isolated by Ficoll separation and stored in liquid nitrogen.

### Kinetics of COVID-19 vaccine mRNA in human blood

A reverse transcription droplet digital PCR (ddPCR) method was developed to detect and quantify COVID-19 vaccine mRNA (see method section for details). All pre-vaccination blood samples (which were a minimum of 139 days after any previous mRNA vaccination, Table S1) were negative for COVID-19 vaccine mRNA (Figure 1b). Vaccine mRNA was detected in the plasma samples of all 19 bivalent booster vaccine subjects at 4 hours post-vaccination (range 6.5–112 mRNA copies µL^-1^, equivalent to 0.005–0.081 ng mL^-1^), peaked at 1–2 (mean 1.3) days post vaccination (at peak levels of up to 731 mRNA copies µL^-1^, equivalent to 0.529 ng mL^-1^), and subsequently displayed log-linear decay kinetics (Figure 1b, Figure S1c). Small amounts of mRNA (0.06–0.6 mRNA copies µL^-1^) remained weakly detectable in 50% of Moderna vaccine subjects’ plasma (9 out of 18 subjects where plasma samples were available) at sday 28 post-vaccination.

### Kinetics of COVID-19 vaccine ionizable lipids in human blood

SM-102 is a non-human ionizable lipid used to interact with mRNA in the lipid nanoparticle formulation of the Moderna SPIKEVAX mRNA vaccine. Since SM-102 has a unique profile separate to human lipids, a mass spectrometry-based lipidomics method was developed to detect and quantify SM-102 lipids in human plasma (Figure 1c, see method section for details). SM-102 background signal was determined in plasma without SM-102 (collected pre-vaccination), and levels were detected above background (range 0.39–8.39 ng mL^-1^) in the plasma of all 19 Moderna vaccine subjects at 4 hours post-vaccination (Figure 1d, Figure S1d). SM-102 levels peaked at 4 hours to 2 days (mean 1.1 day) post-vaccination (median 3.22 ng mL^-1^), and subsequently showed log-linear decay kinetics. The SM-102 signals remained significantly above the background at day 4 post-vaccination (up to 1.16 ng mL^-1^) and approached background levels by day 7 post-vaccination (up to 0.12 ng mL^-1^).

### Degradation on vaccine mRNA in vivo

mRNA is labile in blood at 37 when not protected by a lipid nanoparticle.^15^ Total vaccine mRNA (which is >2000 bp) was detected in plasma in the studies above using a short 113 bp ddPCR reaction, which would detect both intact and some degraded mRNA. To assess the levels of intact and degraded vaccine mRNA in plasma, we adapted a linkage ddPCR technique developed by Hanna et al. which analyses whether both 3’ and 5’ fragments of the vaccine mRNA can be amplified in a single droplet (Figure 2a, see methods section for details).^3^ Amplifying both fragments (double positive events) suggests the mRNA is relatively intact (i.e. spans both PCR reactions), which can be quantified mathematically by comparing single and double-positive event levels. Using this assay, we were able to track the degradation kinetics of the vaccine mRNA in vivo. A representative read-out of the linkage ddPCR assay for 3 timepoints in one subject is shown in Figure 2b. We found the proportion of intact mRNA remains relatively stable within 4–24 hours post-vaccination across 19 subjects (Figure S3). However, the proportion of intact mRNA consistently and slowly fell over time (Figure 2c). Measuring the level of total vaccine mRNA and the proportion of intact vaccine mRNA allowed us to calculate the kinetics of degradation of intact mRNA to non-intact mRNA levels over time, which also show log-linear decay kinetics (Figure 2d).

**Figure 2.**
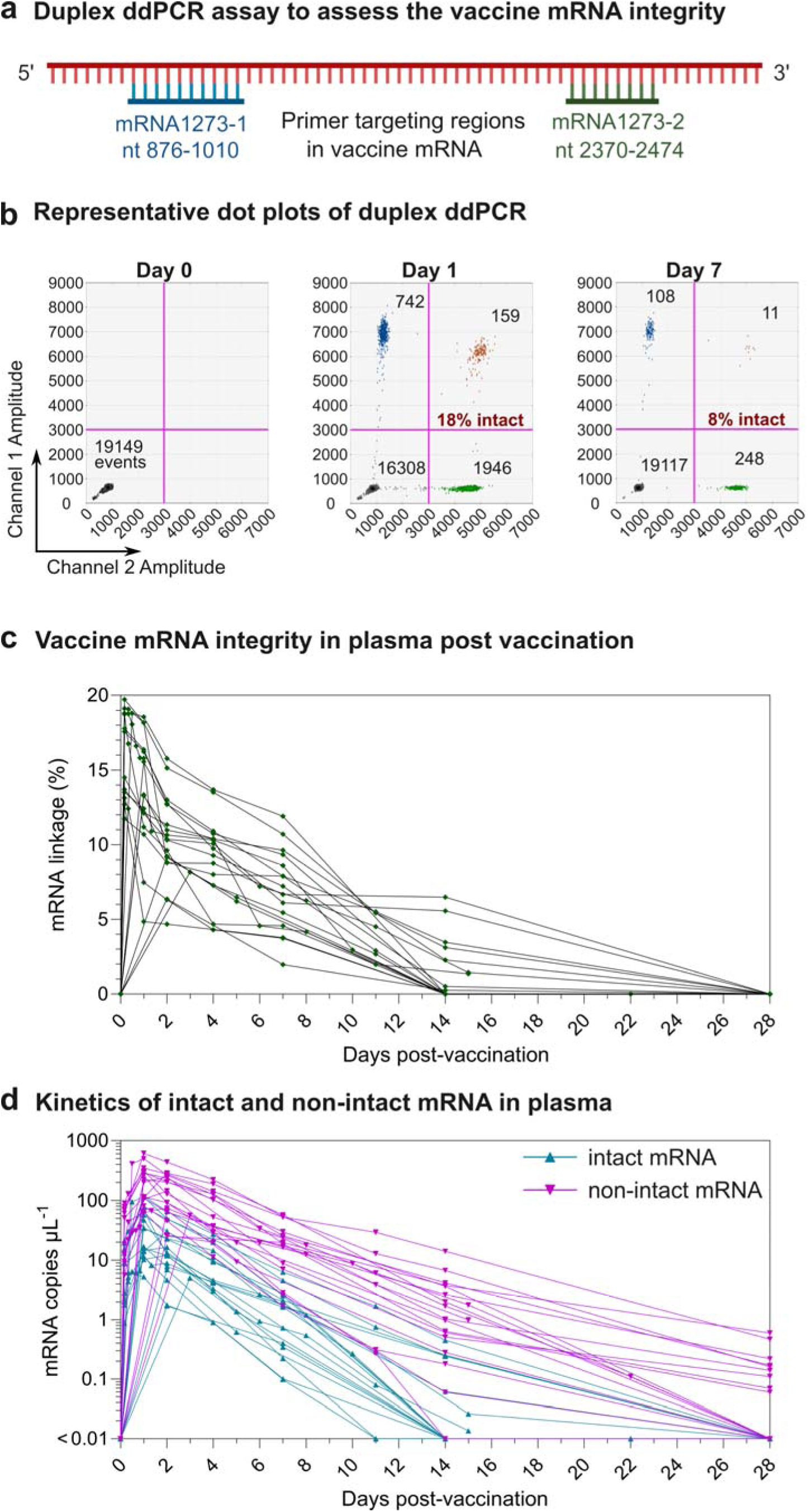
Integrity of vaccine mRNA in plasma after vaccination of SPIKEVAX SARS-CoV-2 mRNA vaccine. **a** Schematic illustration of a duplex ddPCR assay using a two-primer set targeting two regions (mRNA1273-1 nt 876-1010 and mRNA1273-2 nt 2370-2474) of the mRNA1273 sequence. **b** Representative dot plot profiles of FAM-labeled mRNA1273-1 primer and probe (channel 1, amplitude) and HEX-labeled mRNA1273-2 primer and probe (channel 2, amplitude) at day 0 (left panel), 1 (middle panel) and 7 (right panel) post-vaccination. Droplets emitting 2D signals were separated into four groups (Gray: double negative for mRNA1273-1 and mRNA1273-2; Blue: positive for mRNA1273-1, negative for mRNA1273-2; Green: positive for mRNA1273-2, negative for mRNA1273-1; Orange: double positive for both mRNA1273-1 and mRNA1273-2). **c** Vaccine mRNA integrity in plasma of 19 subjects post-vaccination. Vaccine mRNA integrity was assessed by mRNA linkage (%), which was expressed as the estimated percent of linked molecules (correcting for the frequency of random association of probes). The number of droplets in each single or double positive group was derived by QX Manager Software. **d** Longitudinal intact and non-intact mRNA levels in the plasma of the 19 subjects before and after vaccination. The intact mRNA levels were calculated by multiplying the mRNA linkage (%) by the total mRNA levels detected in plasma. To improve readability, the detailed mRNA integrity kinetics within the first 24 hours of vaccination is shown in Figure S3.

The detection of both intact vaccine mRNA and the SM-102 lipid in the blood suggests that the lipid nanoparticle, containing both materials, may be circulating in the blood. If this were the case, the decay kinetics of both elements might be similar. Indeed, the decay rate (mean 0.608 day^-1^) and half-life (1.14 days) of intact mRNA was essentially identical to that of the ionizable lipid (mean decay rate 0.607 day^-1^, half-life 1.14 days, Figure 3). Non-intact mRNA had a slower decay than intact mRNA (half-life 1.43 vs. 1.14 days, Figure 3b). The reason of the longer circulation time of non-intact mRNA is unclear, which is likely to be related with the properties of the lipid nanoparticles when loaded with small fragments of mRNA. The slow degradation of the mRNA despite circulating in blood in vivo at 37 °C (half-life 4.85 days, Figure 3b), and the identical decay rate of intact mRNA and the ionizable lipid, suggests that the mRNA was largely protected in circulation within the lipid nanoparticle.

**Figure 3.**
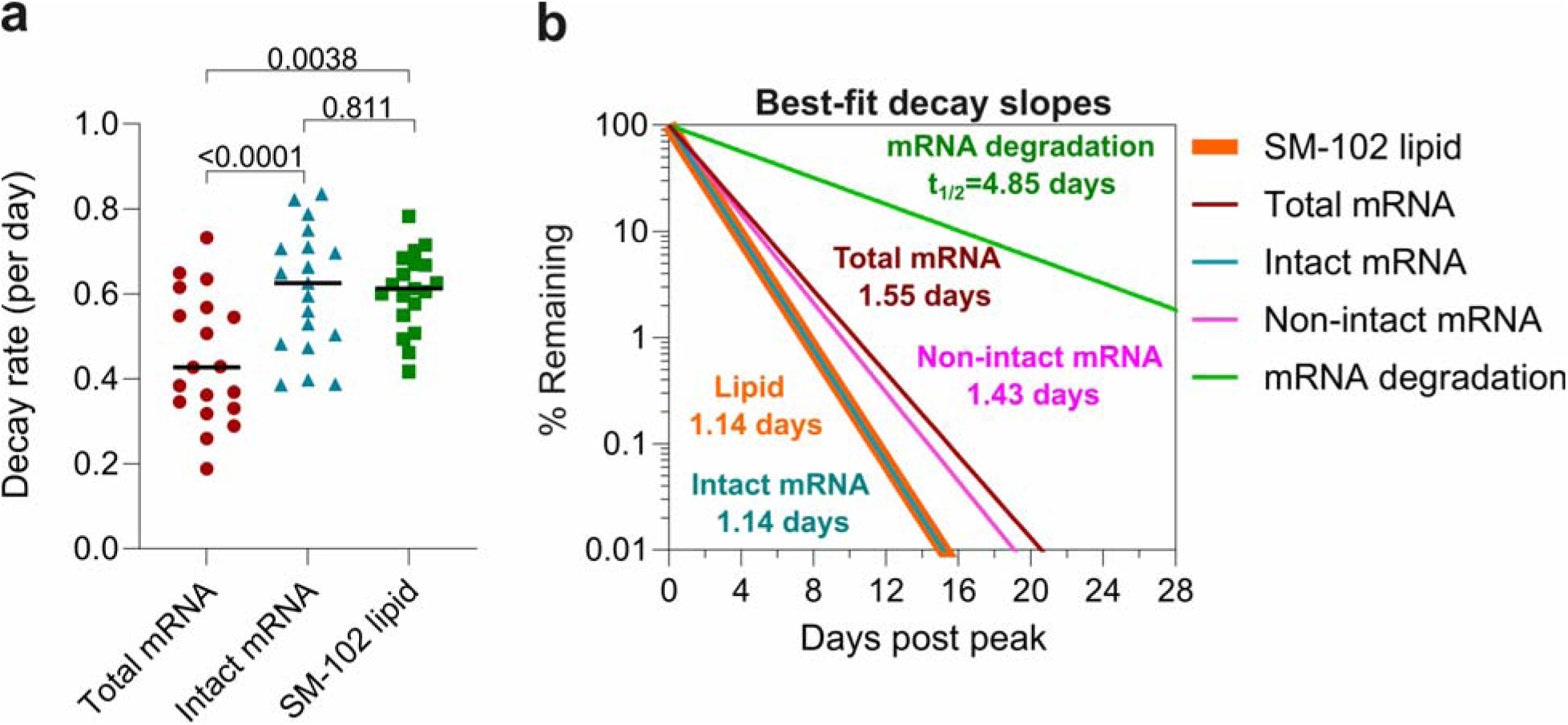
Dynamics of vaccine mRNA and SM-102 lipids in the plasma. **a** Comparison of decay rate between total mRNA, intact mRNA and SM-102 lipid. Statistics assessed by the likelihood ratio test. **b** Best-fit decay slops of SM-102 lipids, total mRNA, intact mRNA, non-intact mRNA, and the rate of degradation of intact mRNA. The responses at the first time point (the peak time) for each parameter are set to 100% and the change (%) over time and half-life are shown. As the decay slopes of SM-102 lipid and intact mRNA overlap, the curve of SM-102 lipid slope was plotted with higher thickness than intact mRNA slope to improve readability.

### Expansion of anti-PEG antibodies

PEG-specific IgG and IgM antibodies in human plasma were quantified by ELISA using our established method.^9^ Anti-PEG IgG and IgM was detectable (end point binding titer >1:10) pre-vaccination in the plasma of 15 and 18 of the 19 subjects, ranging in titer from 1: 15 to 1: 1321 and from 1:26 to 1:1247, respectively (Figure 4a). Following immunization, an increase in PEG-specific IgG and IgM was observed with a mean fold change of 1.4 (range 1–6.7) and 4.6 (range 1–20.3) at day 28 post-vaccination, respectively. This was less than the fold change of PEG IgG (13.1, range 1–70.9) and IgM (68.5, range 0.9– 377.1) following a 2-dose primary Moderna mRNA-1273 vaccination reported in our previous study.^9^ Longitudinal analyses showed PEG-specific antibodies were boosted in a time-dependent manner with a significant increase observed from day 14 post-vaccination (Figure 4a).

**Figure 4.**
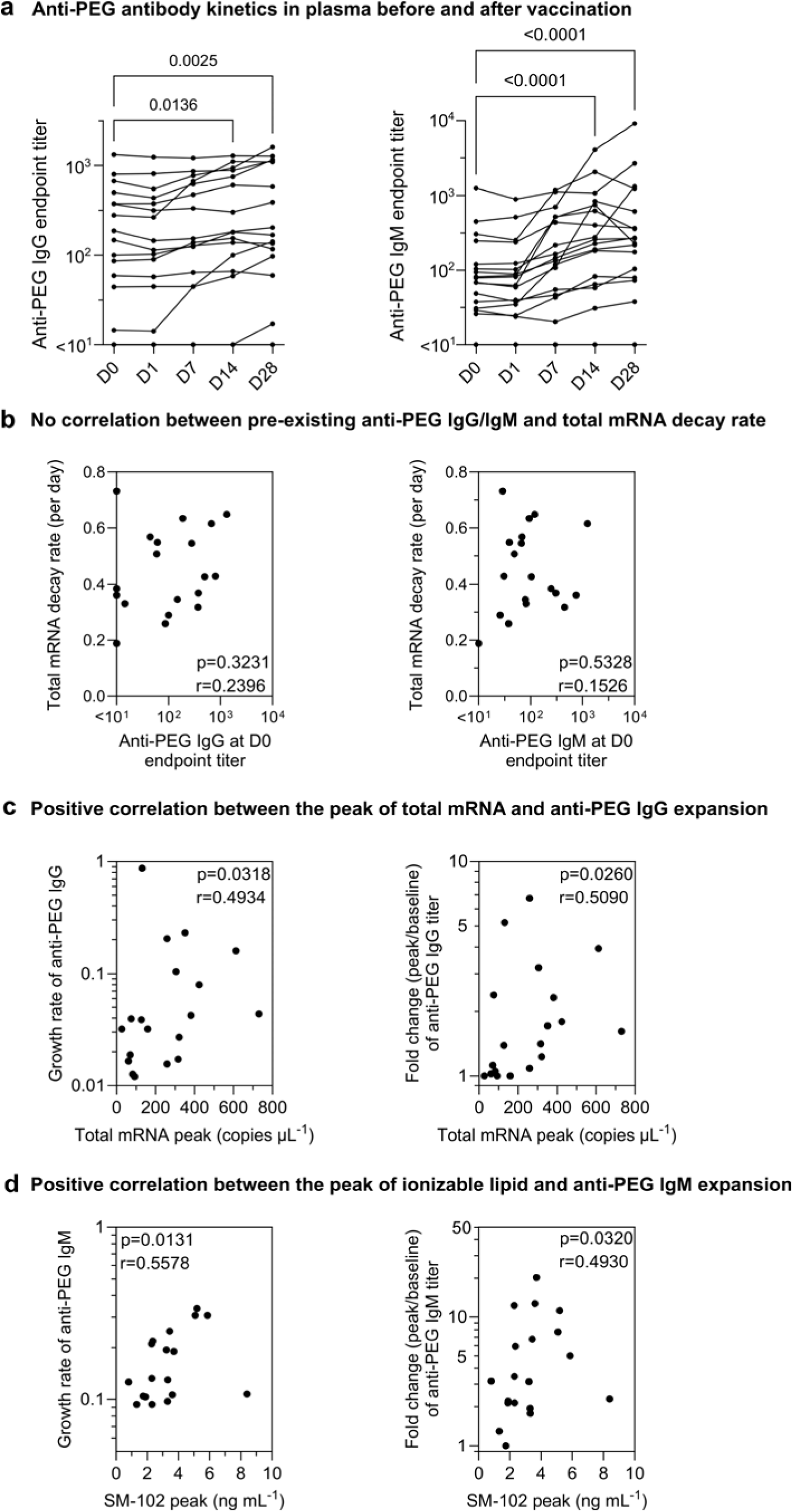
The kinetics of anti-PEG antibody and their correlation with vaccine mRNA kinetics. **a** Longitudinal anti-PEG IgG and IgM titers in the plasma before and after IM inoculation of SPIKEVAX SARS-CoV-2 mRNA vaccine. Statistics assessed by nonparametric Friedman’s test with Dunn’s multiple comparisons test (n=17 as 17 subjects have all five time points). **b** No significant correlation between pre-existing anti-PEG antibody titers and total mRNA decay rate across the 19 subjects. **c** Significant positive correlation between the peak levels of total mRNA in plasma and anti-PEG IgG expansion (growth rate/fold change) across the 19 subjects. **d** Significant positive correlation between the peak levels of ionizable lipid (SM-102) and anti-PEG IgM expansion (growth rate or fold change) across the 19 subjects. Statistics in a-c were assessed by Spearman correlation analysis (n=19).

We hypothesized that the clearance of mRNA lipid nanoparticle might be influenced by the levels of anti-PEG antibodies in the blood through PEG antibody-induced opsonization, a phenomenon called “accelerated blood clearance” phenomenon.^16^ Accelerated clearance of PEGylated nanoparticles by anti-PEG antibodies has been observed in murine studies.^17,18^ However, most murine studies use IV, rather than IM, administration of PEGylated nanoparticles or proteins to induce PEG antibodies and/or detect clearance. The relevance of these IV administration routes in rats or mice to IM administration of mRNA vaccines in humans is unclear.^19^

We found that pre-existing anti-PEG antibodies were not significantly correlated with the decay rate of mRNA or ionizable lipid (Figure 4b, Figure S4). The relatively low levels of mRNA in blood, the relatively uniform decay rates across the 19 subjects, and the generally modest levels of anti-PEG antibodies were potential factors associated with the lack of a correlation between anti-PEG antibodies and mRNA clearance. Interestingly, the peak levels of mRNA and ionizable lipids in the blood post-vaccination positively correlated with subsequent anti-PEG IgG and IgM expansion, respectively (Figure 4c,d), suggesting the boost of anti-PEG antibodies may be influenced by the amounts of lipid nanoparticle mRNA vaccines distributed in the blood.

If anti-PEG antibodies had a major interaction with mRNA lipid nanoparticles in blood, we might expect anti-PEG antibodies to complex with the vaccine-derived nanoparticles and evade detection in our ELISA early after vaccination. However, we found only a very small reduction in anti-PEG IgG antibodies at day 1 post-vaccination compared to pre-vaccination (Figure S5). This suggests that the amounts of lipid nanoparticles distributed into blood were not high enough to have a major impact on the levels of anti-PEG antibodies.

### Expansion of anti-spike and neutralizing antibodies

The vaccine mRNA immunogenicity is reflected by the boost of anti-spike binding and SARS-CoV-2 neutralizing antibodies (Figure 5a,b), which were evaluated by ELISA endpoint dilution titer and a live virus neutralization titer, respectively. All the 19 subjects have received 3 or 4 doses of COVID-19 monovalent vaccination with an interval of 354 (range 139−496) days before the bivalent booster (Table S1). Following the bivalent booster, the spike-specific IgG endpoint titer increased with a mean fold change of 21.3 (range 1.4–302.4) at day 28 post-vaccination in all 19 subjects. As expected, the increase of spike binding antibody titer positively correlated with the boost of neutralizing antibody titer (Figure S6). The neutralizing antibody titer against ancestral (original), BA.1(Omicron subvariant), and BA.5 (Omicron subvariant) strains of SARS-CoV-2 were boosted with a mean fold change of 4.0 (range 0.7–34.9), 15.5 (range 1.1–126.8), and 15.3 (range 1.0–164.7), respectively. The fold change of ancestral neutralization antibodies was lower than BA.1 and BA.5 as all the 19 subjects have already developed a high level of ancestral neutralization antibodies (mean IC_50_ 1838, range 170–34919) prior to the booster vaccine. This is also consistent with the negative correlation between the baseline ancestral neutralizing antibody titer and the spike IgG binding titer increase (Figure 5c). We assessed whether vaccine mRNA immunogenicity was influenced by the levels of mRNA in the blood. However, no correlation between mRNA levels in the blood and the expansion of spike binding IgG or neutralizing antibodies was observed (Figure 5d). The decay rate of mRNA or ionizable lipid was also not influenced by baseline spike binding IgG or neutralizing antibodies (Figure 5e, Figure S7).

**Figure 5.**
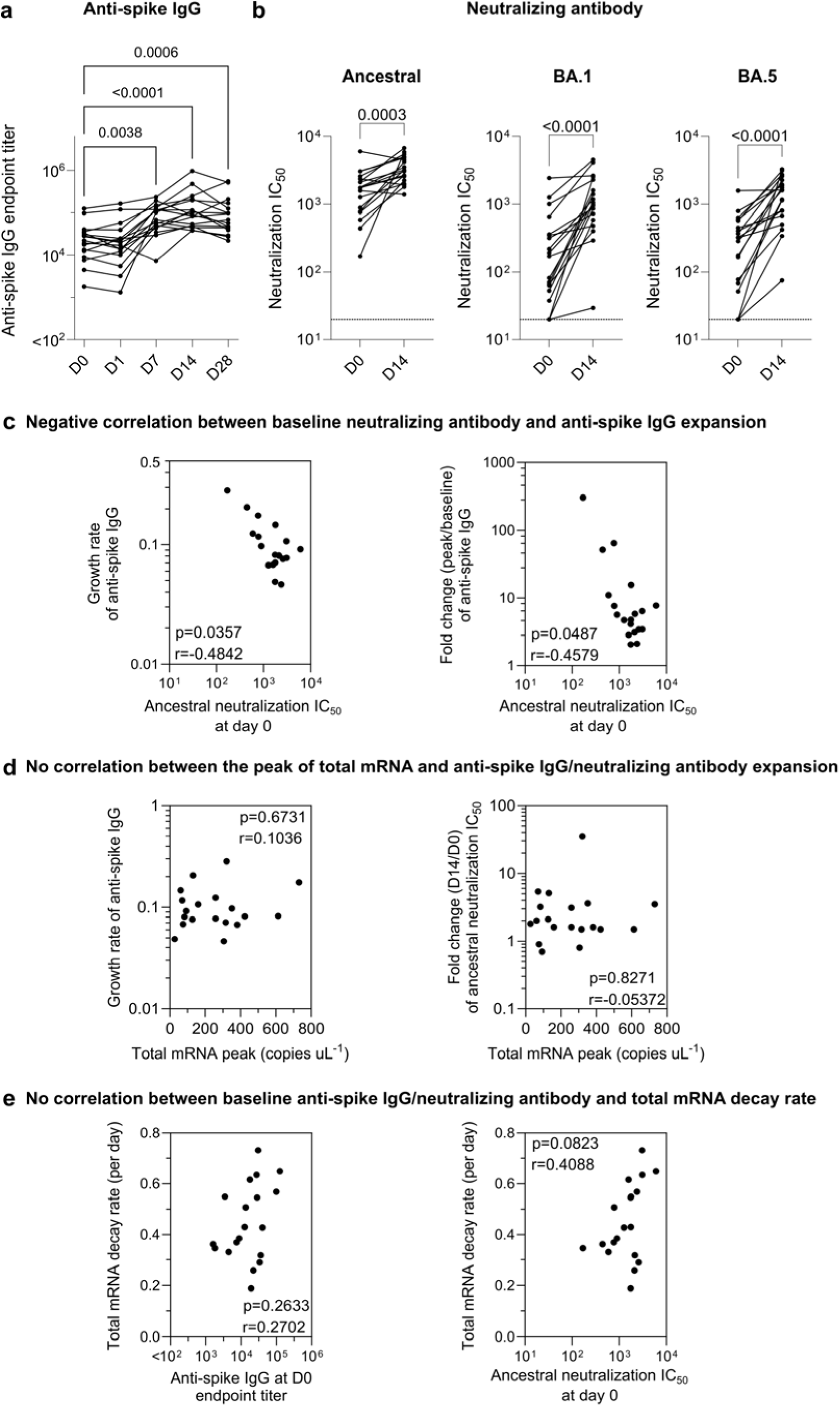
Kinetics of anti-spike IgG and neutralizing antibody. **a** Longitudinal anti-spike IgG titers in the plasma of the Moderna bivalent mRNA vaccinee cohort (Table S1). Statistics assessed by nonparametric Friedman’s test with Dunn’s multiple comparisons test (n=17 as 17 subjects have all five time points). **b** Comparing live SARS-COV-2 neutralization titer (inhibitory concentration 50, IC50) before vaccination (day 0) and post-vaccination at day 14 of the Moderna bivalent mRNA vaccinee cohort. The limit of detection at a titer of 1:20 is shown in the dashed line. Statistics assessed by nonparametric Wilcoxon’s matched-pairs signed rank test (n=19). **c** Significant negative correlation between baseline neutralization antibodies and anti-spike IgG expansion (growth rate or fold change). **d** The peak levels of total vaccine mRNA in the plasma do not influence the expansion of anti-spike IgG and neutralizing antibody. **e** Pre-existing anti-spike IgG and neutralizing antibody do not influence mRNA decay rate. Statistics in c-e were assessed by Spearman correlation analysis.

### The influence of monocyte phagocytosis of lipid nanoparticles

When mRNA lipid nanoparticles are first distributed into the bloodstream, they can be subjected to phagocytosis by blood monocytes and neutrophils. Given the spill over of mRNA vaccines into the blood correlated with anti-PEG antibody expansion (Figure 4c,d), we hypothesized the capacity of blood phagocytes to take up lipid nanoparticles could influence the systemic bioavailability of the lipid nanoparticles and modulate the PEG immunogenicity. Here, we modified a previously developed human blood nanoparticle association assay^20^ to explore the person-specific cellular interactions with primary immune cells (including lymphocytes and monocytes). Peripheral blood mononuclear cells (PBMCs) were available pre-vaccination from 19 of the vaccinees. Cells were washed in serum-free media before incubating with lipid nanoparticles for 1 h at 37 (Figure 6a). The primary immune cells were subsequently labelled with fluorescent antibody cocktails and analysed by flow cytometry to quantify the cellular association of lipid nanoparticles (see details in Methods, gating strategy shown in Figure S8). The lipid nanoparticles were formulated using the same molar composition of lipids as clinically used Moderna (SPIKEVAX) lipid nanoparticle formulation. The lipid nanoparticles displayed a donor-dependent association with monocytes and B cells with minimal association with T cells, natural killer cells, and dendritic cells (Figure 6b and Figure S9). We observed that the ability of monocytes to interact with lipid nanoparticles negatively correlated with the observed increase in anti-PEG IgG titer (Figure 6c, Figure S10), while association with other cell types did not correlate with PEG antibody expansion (Figure S11).

**Figure 6.**
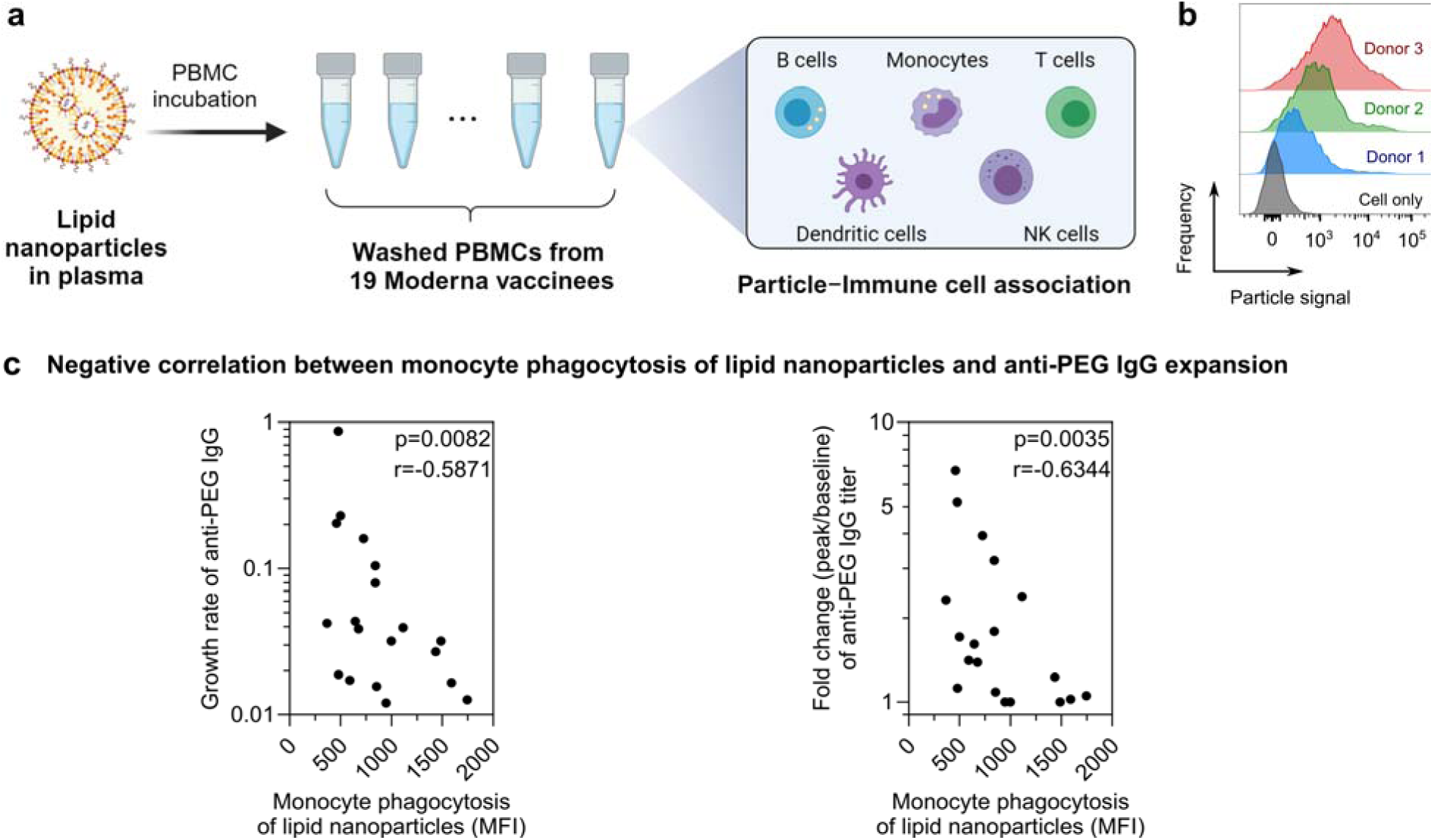
Ex vivo human blood assay to assess lipid nanoparticle−immune cell interactions. Schematic illustration of the in vitro assay to assess the person-specific cellular interactions of primary immune cells with lipid nanoparticles. Peripheral blood mononuclear cells (PBMCs) (collected from the 19 subjects before receiving the Moderna SPIKEVAX bivalent vaccination) were washed by centrifugation with serum-free media multiple times to completely remove plasma. Lipid nanoparticles were pre-incubated with human plasma from one subject and then incubated with PBMCs from the 19 subjects in serum-free media for 1 h at 37 °C, followed by phenotyping cells with antibody cocktails and analysis by flow cytometry. Created with BioRender.com. **b** Flow cytometry histograms represent the monocyte phagocytosis of lipid nanoparticles after incubating with PBMCs of 3 different subjects. Cell only control groups show the respective cell populations without particles in the incubation media. **c** Significant negative correlation between monocyte phagocytosis of lipid nanoparticles (median fluorescence intensity, MFI) and anti-PEG IgG expansion (growth rate or fold change). Statistics were assessed by Spearman correlation analysis.

## CONCLUSIONS

We found that both vaccine mRNA and ionizable lipids can be detected from plasma at 4 hours following the bivalent Moderna SPIKEVAX booster, peak at around day 1 and showed a subsequent log-linear decay profile. We further showed a slow degradation of intact vaccine mRNA in blood. The similar kinetics of intact mRNA and the ionizable lipid in blood and the slow degradation of the mRNA suggests mRNA lipid nanoparticles remain intact and travel from injection sites or lymph nodes into blood stream within 4 hours post-vaccination. The rapid dissemination of mRNA lipid nanoparticles in blood found in our study is consistent with the recent findings on the detection of mRNA in breast milk at 3–45 hours post-vaccination.^3^ We detected low levels of mRNA in plasma 14-28 days after vaccination. This is consistent with recent cross-sectional and autopsy studies.^2,5^ Taken together, our results suggest vaccine mRNA lipid nanoparticles recirculate for up to 1-month post-vaccination.

We initially hypothesised that the decay rate of mRNA (lipid nanoparticles) would be influenced by the levels of anti-PEG antibodies, as many animal studies have demonstrated the phenomenon of accelerated blood clearance.^19^ However, we did not observe such a correlation (Figure 4b). The relatively low levels of anti-PEG antibodies in the blood and relatively uniform levels of mRNA decay across the subjects suggests mRNA decay may be more of an intrinsic feature of humans and less susceptible to external factors such as PEG antibodies. We speculate that humans with much higher levels of PEG antibodies, such as those receiving PEGylated therapeutics intravenously, may clear lipid nanoparticle mRNA vaccines more quickly.

We did however observe that the peak amounts of mRNA and ionizeable lipid (lipid nanoparticles) detected in the blood had a positive correlation with the subsequent expansion of anti-PEG IgG and IgM (Figure 4c,d). We also observed a negative correlation between the level of in vitro monocyte phagocytosis of lipid nanoparticles and anti-PEG IgG expansion (Figure 6c). These findings suggest the amounts of mRNA lipid nanoparticles that remain the blood (and free of phagocytosis) may influence the PEG antibody immunogenicity in humans. This phenomenon only affected PEG immunogenicity, as the mRNA levels in the blood were not significantly correlated with the expansion of spike-binding IgG or neutralizing antibodies (Figure 5d). This is consistent with PEG being expressed on the surface of the lipid nanoparticle, whereas the spike protein, the target of neutralization and spike binding IgG, is only expressed following mRNA transfection of cells.

Additional factors could influence the biodistribution and immunogenicity of mRNA lipid vaccines. Humans display a diverse range of plasma lipids and proteins. Similar differences are likely to be found in lymphatic fluids that vaccines must traverse. The proteins and lipids bind to mRNA lipid nanoparticles in either lymph or plasma, forming so called “biomolecular corona”,^21,22^ which could influence uptake and immunogenicity of the vaccines.^23–26^ We previously showed that clinically relevant drug-loaded liposomes displayed a large variance in immune cell association in human blood, depending on the compositions of person-specific biomolecular coronas.^20^ The formation of biomolecular corona on drug-loaded liposomes *in vivo* in human blood has been previously described.^27^ The composition of biomolecular coronas has been shown to influence the fate and functionality of lipid nanoparticles.^28,29^ An analysis of the in vivo or ex vivo corona formation on mRNA vaccines could help further characterise mRNA vaccine distribution and immunogenicity.

Overall, our study provides important new information on the kinetics of mRNA lipid nanoparticle vaccines in human blood in vivo and their impact on vaccine and PEG immunogenicity. Enhancing our understanding on the biodistribution of mRNA lipid nanoparticles in human should ultimately help improve the safety and efficacy of mRNA vaccines and therapeutics.

## METHODS

### Ethics statement

The study protocols were approved by the University of Melbourne human research and ethics committee (approvals #2056689), and all associated procedures were carried out in accordance with the approved guidelines. All participants provided written informed consent in accordance with the Declaration of Helsinki.

### Participant recruitment and sample collection

Participants were recruited through contacts with the investigators and invited to provide serial blood samples. We recruited 19 participants who received a bivalent Moderna SPIKEVAX booster immunization. Participants characteristics are collated in Table S1. For all participants, whole blood was collected with sodium heparin or ethylenediaminetetraacetic acid (EDTA) anticoagulant or using a serum-separating tube. Plasma and serum were collected and stored at −80 °C, and PBMCs were isolated via Ficoll-Paque separation, cryopreserved in 10% dimethyl sulfoxide (DMSO)/ fetal calf serum (FCS) and stored in liquid nitrogen.

### Quantification of COVID-19 vaccine mRNA

COVID-19 vaccine mRNA was quantified by reverse transcription droplet digital PCR (ddPCR). Total RNA was isolated from 140 µL of plasma (EDTA as anticoagulant) using QIAmp RNA Extraction Kit (Cat# 52906 Qiagen). RNA (10 µL) was subjected to reverse transcription, using Superscript III following the manufacturer’s recommendation (Invitrogen). Based on the putative sequence of Moderna COVID bivalent (SPIKEVAX Bivalent Original/Omicron BA.4-5), the assay was designed and primers and probe were synthesized (Integrated DNA Technologies). These primer and probe set specific to respective codon-modified vaccine mRNA sequence and do not amplify wild-type S-gene (See sequences on Table S2).

Droplet digital PCR was performed using a QX200 Droplet Digital PCR system (Bio-Rad). The ddPCR reaction mixture consisted of 2x ddPCR Supermix (12 µL) for probes (no dUTP, Bio-Rad, cat # 1863024), cDNA (5 µL) and primers/probes mix (5 µL) to a final volume of 24 µL. Droplet generation was achieved using a DG8 Droplet Generator Cartridge (cat # 1864008, Bio-Rad) and Droplet Generator (Bio-Rad). Amplification was carried out on a C1000 Touch thermal cycle (Bio-Rad) using a thermal profile beginning with 95 for 10 min, followed by 40 amplification cycles of 94 for 30 s, and 60 for 60 s, and ending with 98 for 10 min (ramp rate 2 /sec for each step). After PCR, the plate was subsequently read on QX200 droplet reader (Bio-Rad) and data analyzed with QuantaSoft 1.7.4 software.

Preparations incorporating pre-vaccination plasma samples were used as negative controls where there was no vaccine mRNA. Vaccine mRNA was spiked into the negative human plasma (collected pre-vaccination), followed by cDNA reaction, 10-fold series dilution in nuclease-free water and ddPCR reaction to set the positive droplet threshold and generate a linear PCR standard curve. The copy number of the vaccine mRNA in the PCR reaction was used to derive the copy number per mL of plasma. The linear standard curve (Figure S1a) of vaccine mRNA PCR data was derived in GraphPad Prism 10 with relative weighting (weighting by 1/y^2^).

### Quantification of ionizable lipids

Ionizable lipid SM-102 were detected and quantified via targeted mass spectrometry. Lipids were extracted from 50 µL of diluted plasma (90%, a mixture of 5 µL phosphate-buffered saline (PBS) and 45 µL neat plasma) using 500 µL of 1-butanol/methanol (1:1, v/v), followed by vortexing for 10 s and sonication for 60 min in a sonication water bath at 20 . The samples of the linear SM-102 standard curve were prepared by mixing 5 µL of lipid nanoparticles (with known SM-102 concentration in PBS) into 45 µL of plasma (collected pre-vaccination) at a final concentration of SM-102 from 0.1 to 12.5 ng mL^-1^, followed by the same lipid extraction procedure. All the plasma and standard samples were subsequently centrifuged at 16,000 g for 20 mins and 100 µL of the supernatant was transferred to glass vials before mass spectrometry analysis.

Samples were analysed using a Shimadzu 8050 triple quadrupole mass spectrometer coupled to a Shimadzu Nexera X2 liquid chromatography unit. Components of plasma and calibrations samples were separated using an Agilent RRHD Eclipse Plus C18 column (2.1 × 1000 mm, 1.8 μm; Agilent Technologies, USA) over a 15 minute gradient using 6:4 water : acetonitrile containing 10 mM ammonium acetate and 5 µM medronic acid as mobile phase A and 9:1 isopropanol : acetonitrile containing 10 mM ammonium acetate as mobile phase B. The solvent gradient was as follows [time (min), B (%)]: [0, 5], [2, 5], [10, 90], [12, 90], [12.5, 5], [15, 5]. The LC solvent flow rate was 0.3 mL min^-1^ and eluent was diverted to waste for the first 5 minutes of the analysis. The autosampler chamber and column oven were maintained at 10 °C and 40 °C respectively throughout the analysis.

Compounds eluting from the column were introduced into the gas phase by electrospray ionisation (ESI). The nebulising, heating gas flow rate was set to 2 L min^-1^ and both the heating and drying gas flow rates were set to 10 L min^-1^. The interface temperature was set to 350 °C and the interface voltage was 4 kV. SM-102 was analysed in positive ion mode using a multiple reaction monitoring (MRM) strategy (Table S3). Collision energies and parameters for SM-102 detection were optimised using the in-built Shimadzu LabSolutions MRM optimisation tool.

Raw mass spectrometry data was analysed using Skyline (v23.1). Analytes were quantified by integration of the peak areas of the summed MRM transitions and the linear standard curve (Figure S1b) was constructed by weighting of standard sample intensities by 1/x^2^.

### Linkage ddPCR to measure relative levels of intact mRNA

We employed the methods of Hanna et al to perform two simultaneous ddPCR reactions, linkage duplex ddPCR at either end of the Spike mRNA using probes with different fluorescence (FAM or HEX).^3^ Where both PCR tests are positive within a single droplet, this suggests the droplet contains mRNA that spans both regions, that is, is relatively intact. If the droplet contains mRNA for only one end of the mRNA, it suggests the mRNA is relatively degraded. By quantifying the proportion of droplets in which both assays yield amplification, samples containing intact vaccine mRNA (positive linkage) can be distinguished from samples containing fragmented mRNA. The percent of linkage of each sample was expressed as the percentage of linked molecules in relation to the total molecules detected. Linkage number was calculated by Quantasoft 1.7.4 software which determined the excess of double-positive droplets over the expected due to random colocalization of unlinked targets. Following formula is used to calculate the % of Linkage,

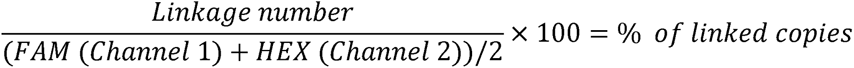

The two sets of probes/primers are shown in Table S4 and the methods are as described in Hanna et al,^3^ which is similar to the ddPCR conditions described for mRNA quantification above.

### Quantification of anti-PEG antibody

The ELISA to detect anti-PEG IgG and IgM was conducted using a previously developed method.^9^ Briefly, the 8-arm PEG-NH_2_ (40 kDa, 200 µg mL^-1^, JenKem Technology, USA) in PBS was coated onto MaxiSorp 96-well plates (Nunc, Denmark) for 18 h at 4 °C, followed by washing with PBS four times. Plates were blocked with 5% (w/v) skim milk powder in PBS for 22 h, followed by adding serially diluted human plasma in 5% skim milk in duplicate for 1 h at 22 °C. Plates were washed with 0.1% 3-[(3-cholamidopropyl)-dimethylammonio]-1-propanesulfonate (CHAPS, Sigma-Aldrich, USA)/PBS buffer twice and PBS four times prior to addition of an HRP-conjugated anti-human IgG (Dako Agilent, USA) at 1:20,000 dilution or HRP-conjugated anti-human IgM (Jackson ImmunoResearch Laboratories, USA) at 1:10,000 for 1 h at 22 °C. Plates were washed as above and then developed using 3,3’,5,5’-Tetramethylbenzidine (TMB) liquid substrate (Sigma-Aldrich, USA). Reaction was stopped with 0.16M H_2_SO_4_ and read at 450nm. Endpoint titres were calculated as the reciprocal plasma dilution giving signal 2× background using a fitted curve (4-parameter log regression) and reported as a mean of duplicates. Background was detected by adding the diluted plasma samples (at 1:10 dilution in 5% skim milk) to the non-PEG-coated wells, followed by the same ELISA procedure.

### Quantification of anti-spike IgG

Plasma antibody binding to SARS-CoV-2 ancestral spike protein was tested by ELISA. 96-well Maxisorp plates were coated overnight at 4 with 2 µg mL^-1^ recombinant spike protein (Hexapro^30^). After blocking with 1% FCS in PBS, duplicate wells of four-fold serially diluted plasma (1:100 – 1:1,638,400) were added and incubated for two hours at room temperature. Bound antibody was detected using 1:20,000 dilution of HRP-conjugated anti-human IgG (Agilent, P021402-5, USA). Plates were then developed using TMB substrate, stopped using sulphuric acid and read at 450nm. Plates were washed 3-6 times with PBS + 0.05% Tween-20 following incubations. Endpoint titres were calculated using Graphpad Prism as the reciprocal serum dilution that gave an OD reading of 2× background using a fitted curve (4 parameter log regression).

### SARS-CoV-2 virus propagation and titration

Ancestral SARS-CoV-2 (hCoV-19/Australia/VIC01/2020) isolate was grown in Vero cells in serum-free DMEM with 1 µg ml^-1^ TPCK trypsin while Omicron BA.1 (hCoV-19/Australia/NSW-RPAH-1933/2021) and BA.5 (hCoV-19/Australia/VIC61194/2022) strains were grown in Calu3 cells in DMEM with 2% FCS. Cell culture supernatants containing infectious virus were harvested on Day 3 for VIC01 and Day 4 for Omicron strains, clarified via centrifugation, filtered through a 0.45µM cellulose acetate filter and stored at -80°C.

Infectivity of virus stocks was then determined by titration on HAT-24 cells (a clone of transduced HEK293T cells stably expressing human ACE2 and TMPRSS2^31,32^). In a 96-well flat bottom plate, virus stocks were serially diluted five-fold (1:5-1:78,125) in DMEM with 5% FCS, added with 30,000 freshly trypsinised HAT-24 cells per well and incubated at 37°C. After 46 hours, 10 µl of alamarBlue™ Cell Viability Reagent (ThermoFisher) was added into each well and incubated at 37 for 1 hour. The reaction was then stopped with 1% SDS and read on a FLUOstar Omega plate reader (excitation wavelength 560nm, emission wavelength 590nm). The relative fluorescent units (RFU) measured were used to calculate %viability (‘sample’ ÷ ’no virus control’ × 100), which was then plotted as a sigmoidal dose response curve on Graphpad Prism to obtain the virus dilution that induces 50% cell death (50% infectious dose; ID_50_). Each virus was titrated in quintuplicate in 3-5 independent experiments to obtain mean ID_50_ values.

### SARS-CoV-2 microneutralisation assay with viability dye readout

Plasma neutralization activity was measured against live SARS-CoV-2 (ancestral, BA.1 and BA.5) as described previously^32^. In 96-well flat bottom plates, heat-inactivated plasma samples were diluted 3-fold (1:20-1:43,740) in duplicate and incubated with SARS-CoV-2 virus at a final concentration of 2× ID_50_ at 37°C for 1 hour. Next, 30,000 freshly trypsinized HAT-24 cells in DMEM with 5% FCS were added and incubated at 37°C. ‘Cells only’ and ‘Virus+Cells’ controls were included to represent 0% and 100% infectivity respectively. After 46 hours, 10µl of alamarBlue™ Cell Viability Reagent (ThermoFisher) was added into each well and incubated at 37°C for 1 hour. The reaction was then stopped with 1% SDS and read on a FLUOstar Omega plate reader (excitation wavelength 560nm, emission wavelength 590nm). The relative fluorescent units (RFU) measured were used to calculate %neutralisation with the following formula: (‘Sample’ – ‘Virus+Cells’) ÷ (‘Cells only’ – ‘Virus+Cells’) × 100. Inhibitory concentration 50 (IC_50_) values were determined using four-parameter non-linear regression in GraphPad Prism with curve fits constrained to have a minimum of 0% and maximum of 100% neutralisation.

### Lipid nanoparticle preparation and characterization

Lipid nanoparticles were formulated with heptadecan-9-yl 8-[2-hydroxyethyl-(6-oxo-6-undecoxyhexyl)amino]octanoate (SM-102, MedChemExpress, USA), distearoylphosphatidylcholine (DSPC, Avanti Polar Lipids, USA), cholesterol (Sigma-Aldrich, USA), and 1,2-dimyristoyl-rac-glycero-3-methoxypolyethylene glycol-2000 (PEG2000-DMG) (MedChemExpress, USA) with the same molar composition of lipids used in the US FDA-approved Moderna SPIKEVAX formulation, in addition to 0.1 mol % dioctadecyl-3,3,3,3-tetramethylindodicarbocyanine (DiD, Thermo Fisher Scientific, USA), using the NanoAssemblr platform (Precision NanoSystems, Canada). Particles were loaded with a nonimmunogenic nucleic acid cargo (firefly luciferase plasmid DNA, PlasmidFactory GmbH & Co. KG, Germany) in order to regulate lipid packing and particle size. The concentration of encapsulated pDNA in lipid nanoparticles (98 μg mL^-1^) was determined using the Quant-iT™ PicoGreen™ dsDNA Assay kit with the encapsulation efficiency >97%. The size (68 ± 18 nm) and polydispersity index (0.08) of the lipid nanoparticles were determined by dynamic light scattering (DLS) analysis performed on a Zetasizer Nano-ZS instrument (Malvern Instruments, UK). The zeta-potential (-10 ± 3 mV) of the lipid nanoparticles were determined by using a Zetasizer Nano-ZS (Malvern Instruments, UK) where particles were dispersed at pH 7.4 in phosphate buffer (5 mM).

### Blood assay to determine lipid nanoparticle association with human immune cells

The frozen PBMCs from the 19 subjects (collected before receiving the Moderna SPIKEVAX vaccination) were thawed at 37, washed twice with serum-free RPMI 1640 medium, and counted with a Cell-DYN Emerald analyzer. The DiD-labelled nanoparticles (3.4 µg based on pDNA loading) were preincubated in 680 µL of plasma (collected from 1 subject pre-vaccination) at 37 for 1 h. The lipid nanoparticles (0.05 µg) in the presence of plasma were subsequently incubated with PBMCs (5 × 10^5^) from 19 subjects in the serum-free RPMI 1640 medium (100 µL) at a particle concentration of 0.45 µg mL^−1^ for 1 h at 37 . After incubation, PBMCs were washed with PBS (4 mL, 500g, 7 min) and stained for phenotypic markers in PBS at 4 °C for 1 h using titrated concentration of antibodies against CD45 V500 (H130, BD), CD19 BV650 (HIB19, BioLegend), CD14 (MΦP9, BD), CD3 AF700 (SP34-2, BD), CD56 PE (B159, BD), lineage-1 (Lin-1) cocktail FITC (BD), and HLA-DR PerCP-Cy5.5 (G46-6, BD). Unbound antibodies were removed by washing twice with cold (4 °C) PBS containing 0.5% w/v BSA and 2 mM EDTA (4 mL, 500 g, 7 min). Cells were fixed with 1% w/v formaldehyde in PBS and directly analyzed by flow cytometry (LSRFortessa, BD Bioscience). The data were processed using FlowJo V10 with the gating strategy shown in Figure S8.

### Estimating the decay rates

The decay rate of mRNA and lipid in plasma was estimated by fitting a linear mixed effect model as a function of days post-peak and response type (mRNA vs lipid). Likelihood ratio test was used to determine if the decay rate is different with respect to the response type. We fitted the model to log-transformed data of various response variables (assuming exponential decay), and we censored the data from below (left-censoring) if it was less than the lower limit of quantitation. The model was fitted by using *lmec* library in *R (v4*.*2*.*1)*, using the maximum likelihood algorithm.

The activation time and growth rate of PEG IgG, PEG IgM, and Spike IgM following vaccination was estimated as previously described.^33^ A piecewise model was used in which the immune response *y* for subject *i* at time y_i_ can be written as:

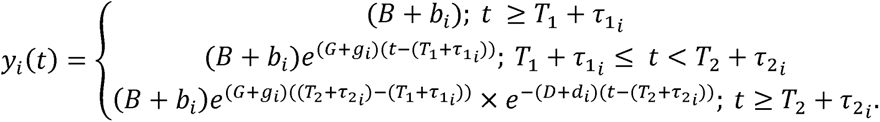

The model has 5 parameters; *B,G,T*_1_*,D*. For a period before *T*_2_, we assumed a constant baseline value *B*, for the immune response (which is higher than or at the background level). After the activation time *T*_1_, the immune response will grow at a rate of G. until *T*_2_. From *T*_2_, the immune response will decay at a rate of *D*. For each subject *i*, the parameters were taken from a normal distribution, with each parameter having its own mean (fixed effect). A diagonal random effect structure was used, where we assumed there was no correlation within the random effects. The model was fitted to the log-transformed data values, with a constant error model distributed around zero with a standard deviation σ. To account for the values less than the limit of detection, a censored mixed effect regression was used to fit the model. Model fitting was performed using Monolix2023R1.

### Statistical analysis

Associations between mRNA, anti-PEG antibodies, anti-spike IgG, neutralizing antibodies and cellular association of lipid nanoparticles were assessed using nonparametric Spearman correlation in GraphPad Prism 10. Decay rates of total mRNA, intact mRNA, and SM-102 lipid were compared by the likelihood ratio test in *R (v4*.*2*.*1)*. Pair-wise comparison of neutralization IC_50_ between D0 and D14 and anti-PEG antibody between D0 and D1 were assessed by nonparametric Wilcoxon’s matched-pairs signed rank test in GraphPad Prism 10. Longitudinal comparison in anti-PEG antibodies, anti-spike IgG and SM-102 signals were derived by nonparametric Friedman’s test with Dunn’s multiple comparisons test in GraphPad Prism 10.

## Data Availability

All data produced in the present study are available upon reasonable request to the authors

## ASSOCIATED CONTENT

**Supporting Information.** Demographic information of donors who received the Moderna SPIKEVAX SARS-CoV-2 bivalent mRNA vaccine; sequences of primer and probe designed for PCR assay; optimised MRM parameters for SM-102 detection; standard curves and concentration of mRNA and ionizable lipids in plasma; kinetics and integrity of vaccine mRNA within first 24 h post-vaccination; correlation between pre-existing anti-PEG antibodies and the decay rate of ionizable lipid; anti-PEG antibody levels pre-vaccination and day 1 post-vaccination; correlation between the boost of spike IgG and the boost of neutralization antibody; correlation between pre-existing anti-spike IgG/neutralizing antibody and the decay rate of ionizable lipid; gating strategy used to identify immune cell populations from human PBMCs; influence of person-specific PBMCs on lipid nanoparticle−immune cell interactions; correlation between monocyte phagocytosis of lipid nanoparticles and anti-PEG IgG expansion; correlation between B cell/T cell/NK cell/Dendritic cell association (MFI) and anti-PEG IgG expansion.

## Author Contributions

SJK and YJ conceived, designed, and supervised the study and drafted the manuscript. SJK, SL, THA, AR, WSL, MGL, JAJ, AKW, MPD, and YJ performed experiments, analysed the experiment data, and provided technical advice. SJK, THA, JN, HEK, JAJ, AKW and YJ recruited subjects and processed their blood samples. DHO and FC provided intellectual inputs and reagents. All authors approved the final version of the manuscript.

## Notes

The authors declare no competing financial interest.

## ACKNOWLEDGMENT

We thank the participations for the generous involvement and provision of samples. We thank C.-J, Kim and S. Ye (University of Melbourne) for excellent technical assistance and helpful discussion. We acknowledge the Melbourne Mass Spectrometry and Proteomics Facility for provision of lipidomics services. This study was supported by the Australian Research Council (ARC) Discovery Project (DP210103114 to FC, SJK and YJ), the National Health and Medical Research Council (NHMRC) program grant (GNT1149990 to SJK and MPD), the Victorian Critical Vaccinees Collection COVID-19 Research Seed Funding Grant (YJ), an ARC Discovery Early Career Researcher Award (DE230101542 to YJ), and NHMRC Investigator grants (SJK, WSL, GNT2016732 to FC, JAJ, AKW, and MPD). For the purposes of open access, the author has applied a CC BY public copyright license to any Author Accepted Manuscript version arising from this submission.

## Table of Contents graphic

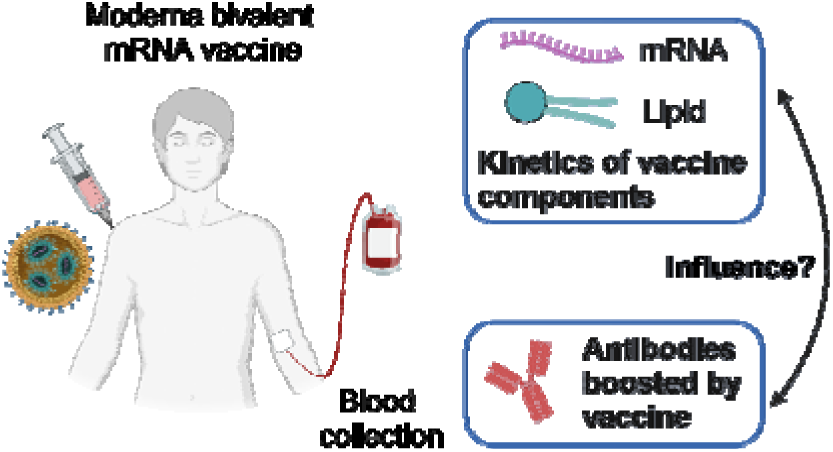

