## Supporting Information for "Blood Distribution of SARS-CoV-2 Lipid Nanoparticle mRNA Vaccine in Humans"

**Table S1.** Demographic information and COVID-19 immunization history of 19 subjects who received the Moderna SPIKEVAX SARS-CoV-2 bivalent mRNA vaccine

| Subject ID | Age | Sex | Vaccine history* | Moderna bivalent vaccination | | |
|---|---|---|---|---|---|---|
| | | | | Type | Dose | Days since last vaccination |
| 1 | 36−40 | M | Pfizer 3 dose | Ancestral & BA.1 | $4^{th}$ | 401 |
| 2 | 26−30 | F | Pfizer 3 dose | Ancestral & BA.1 | $4^{th}$ | 343 |
| 3 | 31−35 | F | Pfizer 3 dose | Ancestral & BA.1 | $4^{th}$ | 424 |
| 4 | 36−40 | M | AstraZeneca 2 dose + Moderna 1 dose + Pfizer 1 dose | Ancestral & BA.1 | $5^{th}$ | 191 |
| 5 | 61−65 | M | AstraZeneca 2 dose + Moderna 1 dose + Novavax 1 dose | Ancestral & BA.1 | $5^{th}$ | 192 |
| 6 | 26−30 | F | Pfizer 3 dose | Ancestral & BA.1 | $4^{th}$ | 346 |
| 7 | 56−60 | F | AstraZeneca 2 dose + Moderna 1 dose | Ancestral & BA.5 | $4^{th}$ | 412 |
| 8 | 61−65 | F | AstraZeneca 2 dose + Pfizer 2 dose | Ancestral & BA.5 | $5^{th}$ | 256 |
| 9 | 56−60 | F | AstraZeneca 2 dose + Moderna 1 dose | Ancestral & BA.5 | $4^{th}$ | 486 |
| 10 | 21−25 | F | AstraZeneca 2 dose + Moderna 2 dose | Ancestral & BA.5 | $5^{th}$ | 139 |
| 11 | 61−65 | M | Novavax 1 dose + Pfizer 3 dose | Ancestral & BA.5 | $5^{th}$ | 270 |
| 12 | 31−35 | F | Pfizer 2 dose + Moderna 1 dose | Ancestral & BA.5 | $4^{th}$ | 432 |
| 13 | 31−35 | M | Pfizer 3 dose | Ancestral & BA.5 | $4^{th}$ | 433 |
| 14 | 26−30 | F | Pfizer 3 dose | Ancestral & BA.5 | $4^{th}$ | 433 |
| 15 | 21−25 | F | AstraZeneca 3 dose | Ancestral & BA.5 | $4^{th}$ | 419 |
| 16 | 26−30 | M | Pfizer 3 dose | Ancestral & BA.5 | $4^{th}$ | 457 |
| 17 | 31−35 | F | Pfizer 1 dose + Moderna 2 dose | Ancestral & BA.5 | $4^{th}$ | 224 |
| 18 | 61−65 | F | AstraZeneca 2 dose + Moderna 1 dose | Ancestral & BA.5 | $4^{th}$ | 496 |
| 19 | 66−70 | M | AstraZeneca 2 dose + Moderna 1 dose | Ancestral & BA.5 | $4^{th}$ | 380 |
| Summary | Mean 42 (range 24−70) | F 63% | Median 3 vaccines *Note: all the vaccines above are monovalent | BA.1: n=6 BA.5: n=13 | $4^{th}$ 74%; $5^{th}$ 26% | Mean 354 (range 139−496) |

**Table S2.** Sequences of primer and probe designed specific to respective codon-modified vaccine mRNA sequence for ddPCR to quantify the levels of vaccine mRNA in plasma

| Primer or probe | Vaccine | Sequence |
|---|---|---|
| AMmRNA-F | mRNA-1273 | 5-'GAGCCTGCTGATCGTGAATAA-3' |
| AMmRNA-P | mRNA-1273 | 5'-TCAAGGTGTGCGAGTTCCAGTTCT-3' |
| AMmRNA-R | mRNA-1273 | 5'-TCCAGCTCTTGTTGTTCTTGT-3' |

F-forward primer, P-probe, R-reverse primer.

**Table S3.** Optimised MRM parameters for SM-102 detection in plasma by mass spectrometry

| Precursor m/z | Product m/z | Q1 pre-bias | Collision energy | Q3 pre-bias | Dwell time (ms) |
|---|---|---|---|---|---|
| 710.6 | 472.4 | -40 | -38 | -23 | 50 |
| 710.6 | 454.4 | -40 | -45 | -24 | 50 |
| 710.6 | 318.2 | -28 | -50 | -12 | 50 |
| 710.6 | 300.2 | -40 | -48 | -20 | 50 |

**Table S4.** Sequences of primer and probe designed specific to respective codon-modified vaccine mRNA sequence for linkage ddPCR to quantify the integrity of vaccine mRNA in plasma

| Primer or probe | Vaccine | Sequence |
|---|---|---|
| mRNA1273-1-F | mRNA-1273 | 5'-GACCTTCCTGCTGAAGTACAA-3' |
| mRNA1273-1-P | mRNA-1273 | 5'-/56-FAM/ACCAAGTGC/ZEN/ACCCTGAAGAGCTTC/3IABkFQ/-3' |
| mRNA1273-1-R | mRNA-1273 | 5'-AAGTTGCTGGTCTGGTAGATG-3' |
| mRNA1273-2-F | mRNA-1273 | 5'-GTGGAGCAGGACAAGAACA-3' |
| mRNA1273-2-P | mRNA-1273 | 5'-/56-HEX/CGCCCAGGT/ZEN/GAAGCAGATCTACAA/3IBkFQ/-3' |
| mRNA1273-2-R | mRNA-1273 | 5'-AGGATCTGGCTGAAGTTGAAG-3' |

F-forward primer, P-probe, R-reverse primer.

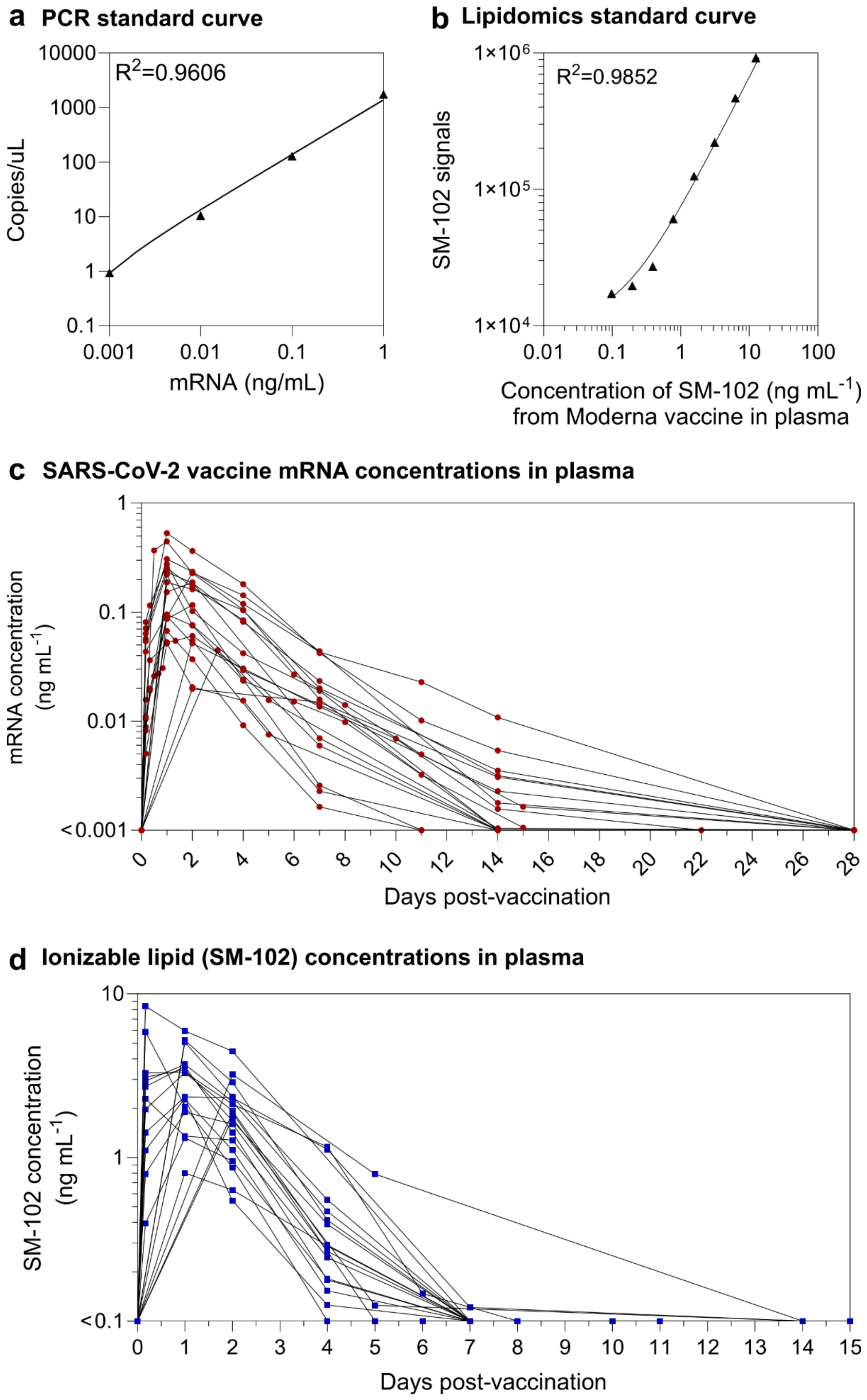

a PCR standard curve
$R^2$=0.9606
10000
1000
100
10
1
0.1
Copies/uL
0.001
0.01
0.1
1
mRNA (ng/mL)
b Lipidomics standard curve
$R^2$=0.9852
1×10^6
1×10^5
1×10^4
SM-102 signals
0.01
0.1
1
10
100
Concentration of SM-102 (ng mL^-1)
from Moderna vaccine in plasma
c SARS-CoV-2 vaccine mRNA concentrations in plasma
1
0.1
0.01
<0.001
mRNA concentration
(ng mL^-1)
0 2 4 6 8 10 12 14 16 18 20 22 24 26 28
Days post-vaccination
d Ionizable lipid (SM-102) concentrations in plasma
10
1
<0.1
SM-102 concentration
(ng mL^-1)
0 1 2 3 4 5 6 7 8 9 10 11 12 13 14 15
Days post-vaccination

**Figure S1.** Converting the raw PCR and lipidomics signals to vaccine mRNA and ionizable lipid (SM-102) concentrations in the plasma. (a) The PCR standard curve of SARS-CoV-2 vaccine mRNA in human plasma, which was used to convert raw PCR signals (see Fig. 1b) to mRNA concentrations (Figure S1c). Vaccine mRNA was spiked into the negative human plasma (collected pre-vaccination), followed by cDNA reaction, 10-fold series dilution in nuclease-free water and ddPCR reaction to generate a linear PCR standard curve. (b) The lipidomics standard curve of SM-102 in human plasma, which was used to convert raw SM-102 signals (see Fig. 1d) to SM-102 concentrations (Figure S1d). The samples of the linear lipidomics standard curve were prepared by titrating the lipid nanoparticles (with known SM-102 concentration in PBS) into the human plasma (from one donor collected pre-vaccination) at different concentration of SM-102 (from 0.1 to 12.5 ng $mL^{-1}$). (c,d) Concentration kinetics of SARS-CoV-2 vaccine mRNA and SM-102 in the plasma of the 19 subjects who received one dose of Moderna SPIKEVAX bivalent mRNA vaccine (see Table S1 for subject details). The lower limit of quantification (LLOQ) is 0.001 and 0.1 ng $mL^{-1}$ for mRNA and SM-102 concentration based on their linear standard curves, respectively.

**a Raw PCR data of vaccine mRNA in plasma within 24 h post-vaccination**

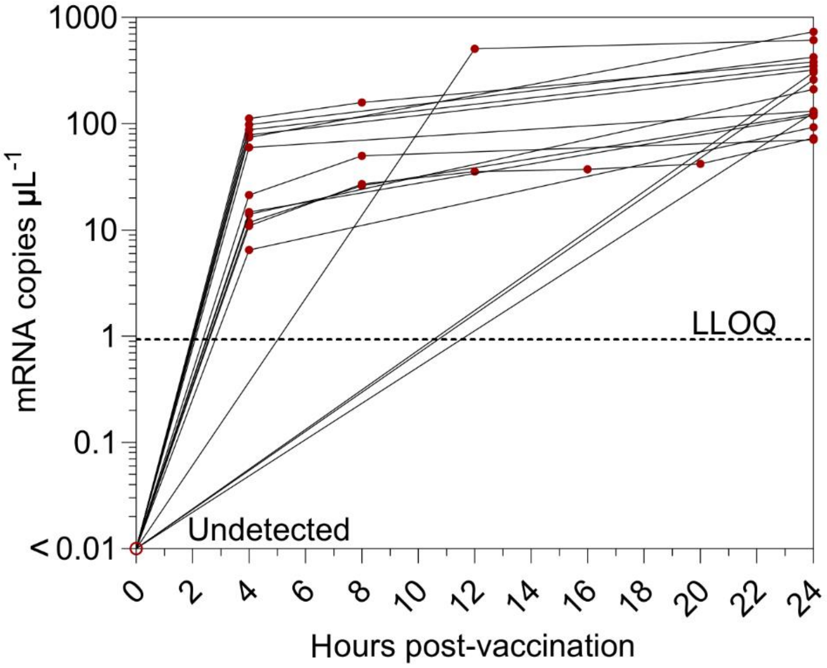


**b Concentrations of vaccine mRNA in plasma within 24 h post-vaccination**

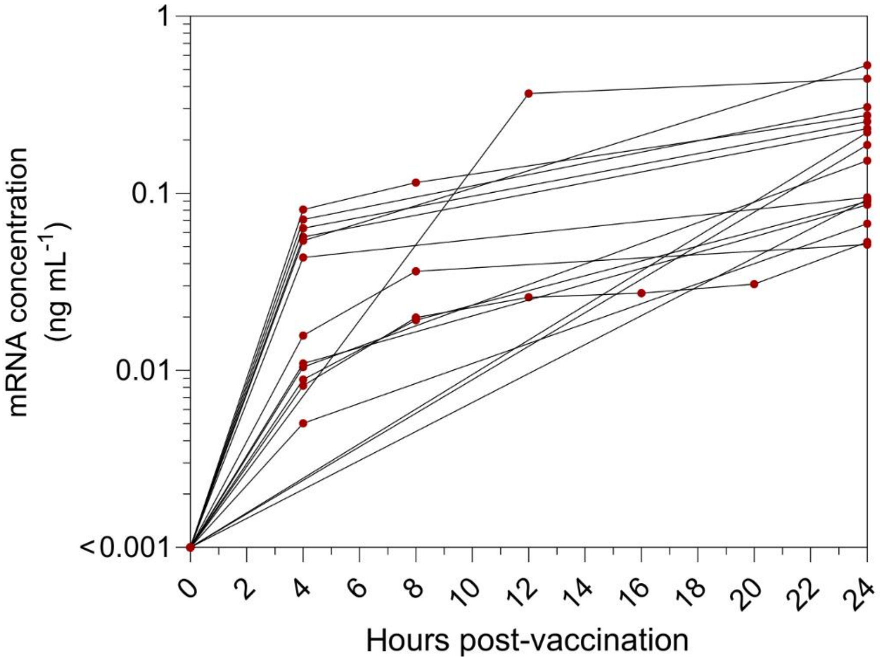


**Figure S2.** Kinetics of SARS-CoV-2 vaccine mRNA in the plasma of the 19 subjects in the first 24 hours post-vaccination of the SPIKEVAX SARS-CoV-2 mRNA vaccine. (a) Longitudinal mRNA levels in the plasma of the 19 subjects before and after receiving one dose of Moderna

SPIKEVAX COVID-19 bivalent mRNA vaccine. LLOQ is shown as a dashed line and the undetected samples were plotted at <0.01 copies $\mu L^{-1}$ with open red circles in the raw PCR data plot. (b) Concentration of vaccine mRNA in the plasma of the 19 subjects before and after receiving one dose of Moderna SPIKEVAX COVID-19 bivalent mRNA vaccine. LLOQ is 0.001 ng $mL^{-1}$ for mRNA concentration based on its linear standard curve.

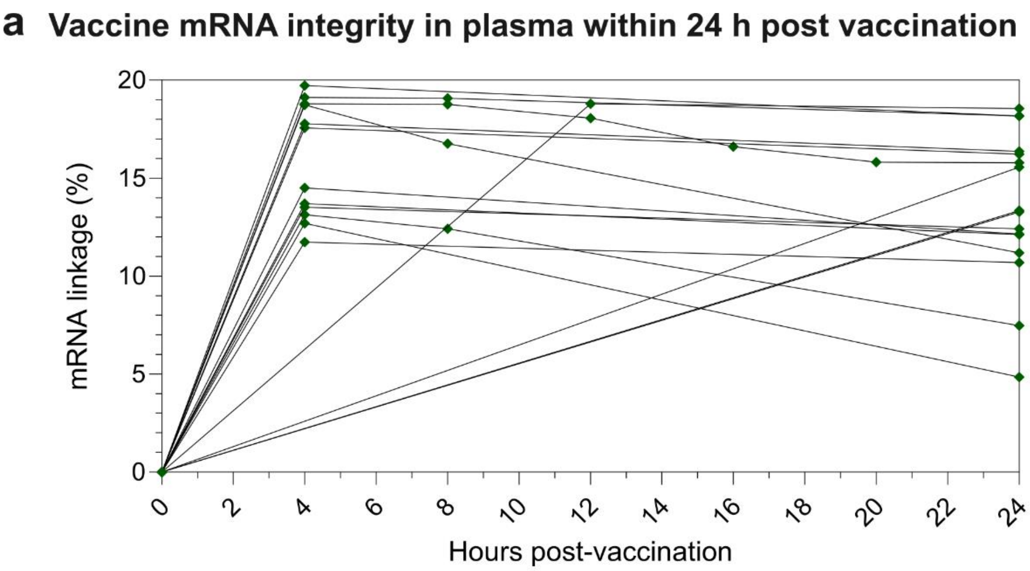


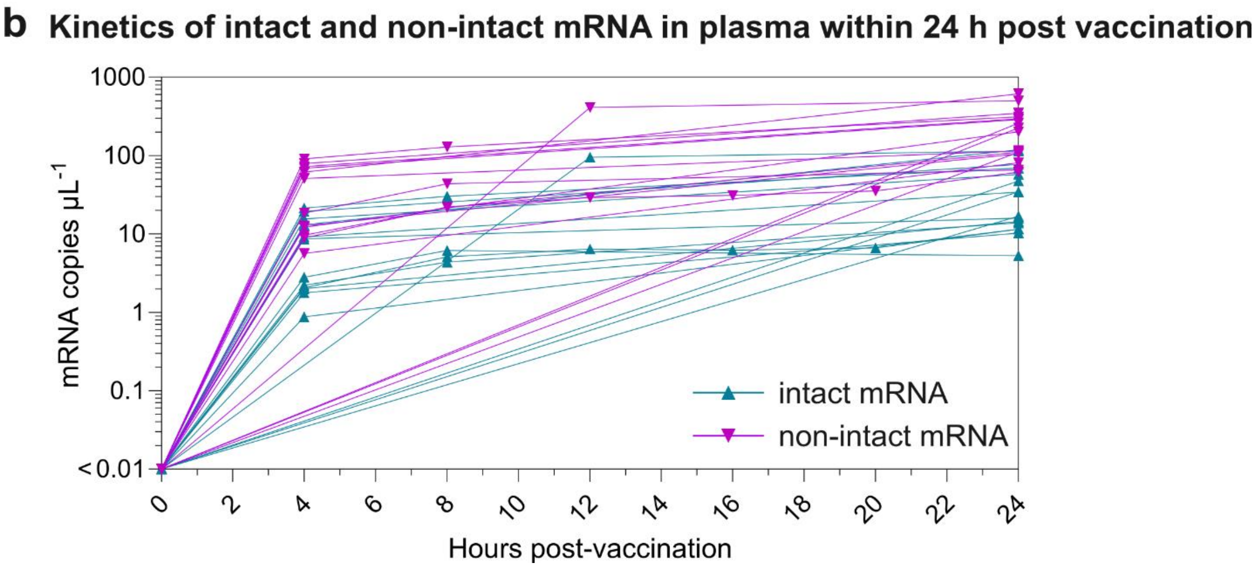


**Figure S3.** Integrity and kinetics of SARS-CoV-2 vaccine mRNA in the plasma of the 19 subjects within 24 h post-vaccination of the SPIKEVAX SARS-CoV-2 mRNA vaccine. (a) Vaccine mRNA integrity in plasma of 19 subjects within 24 h post-vaccination. Vaccine mRNA integrity was assessed by mRNA linkage (%), which was expressed as the estimated percent of linked molecules (correcting for the frequency of random association of probes). The number of droplets in each single or double positive group was derived by QX Manager Software. (b) Longitudinal kinetics of intact and non-intact mRNA levels in the plasma of the 19 subjects within 24 h post-vaccination.

The intact mRNA levels were calculated by multiplying the mRNA linkage (%) by the total mRNA levels detected in plasma.

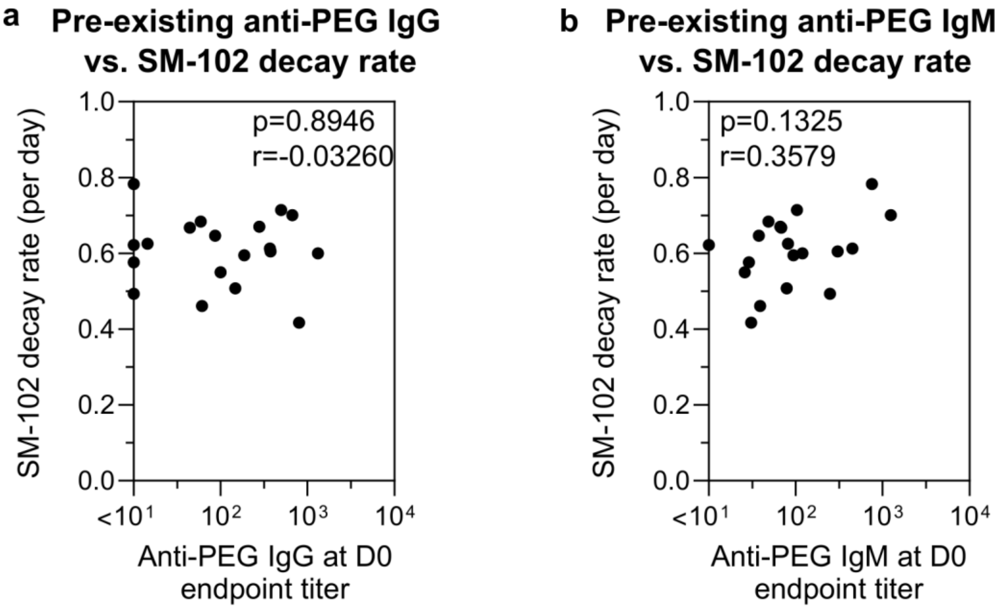


**Figure S4.** Baseline levels of anti-PEG antibodies do not influence the ionizable lipid decay rate in the plasma post-vaccination. (a) No significant correlation between pre-existing anti-PEG IgG and the SM-102 decay rate. (b) No significant correlation between pre-existing anti-PEG IgM and the SM-102 decay rate. Statistics were assessed by Spearman correlation analysis.

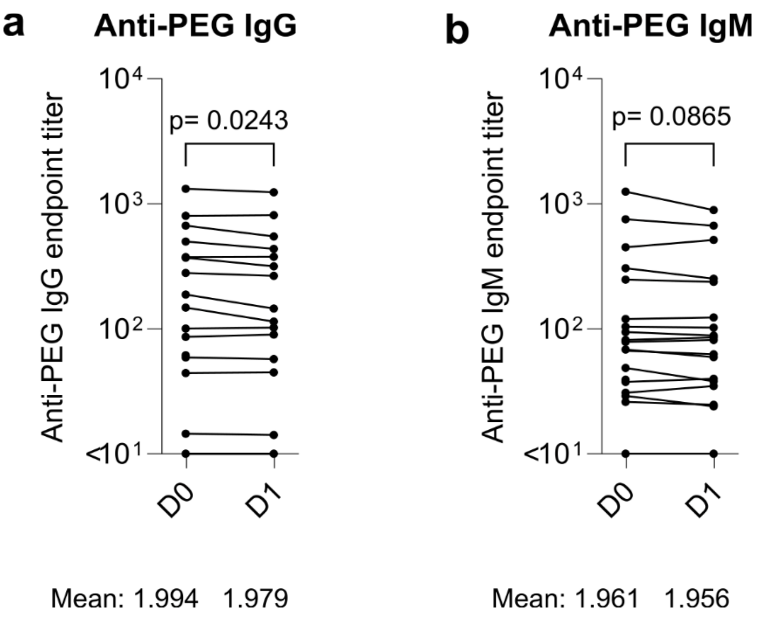


**Figure S5.** Anti-PEG antibodies pre-vaccination and day 1 post-vaccination. Comparing anti-PEG (a) IgG and (b) IgM titers in the plasma of Moderna bivalent mRNA vaccinee cohort at D0 (pre-vaccination) and day 1 post vaccination (D1). Statistics assessed by nonparametric Wilcoxon's matched-pairs signed rank test.

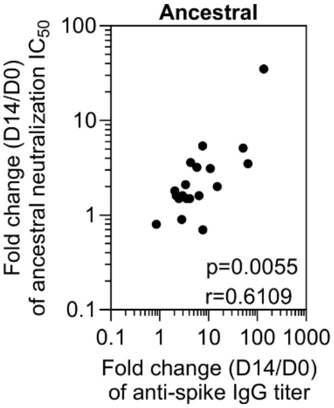


**Figure S6.** Significant positive correlation between the boost of spike IgG and the boost of neutralization antibody. Statistics were assessed by Spearman correlation analysis.

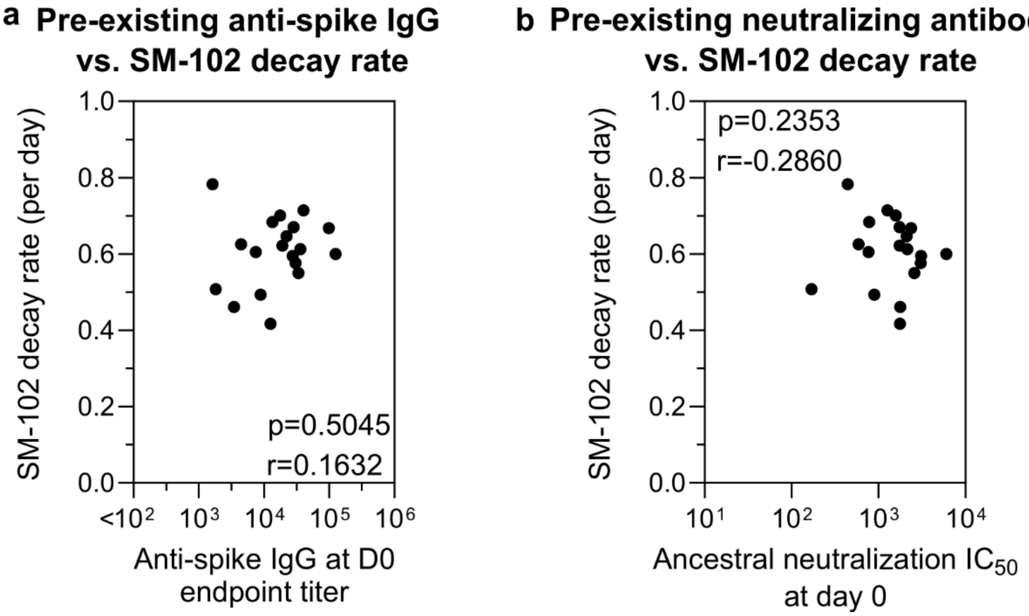


**Figure S7.** Pre-existing anti-spike IgG and neutralizing antibody do not influence ionizable lipid decay rate. Statistics were assessed by Spearman correlation analysis.

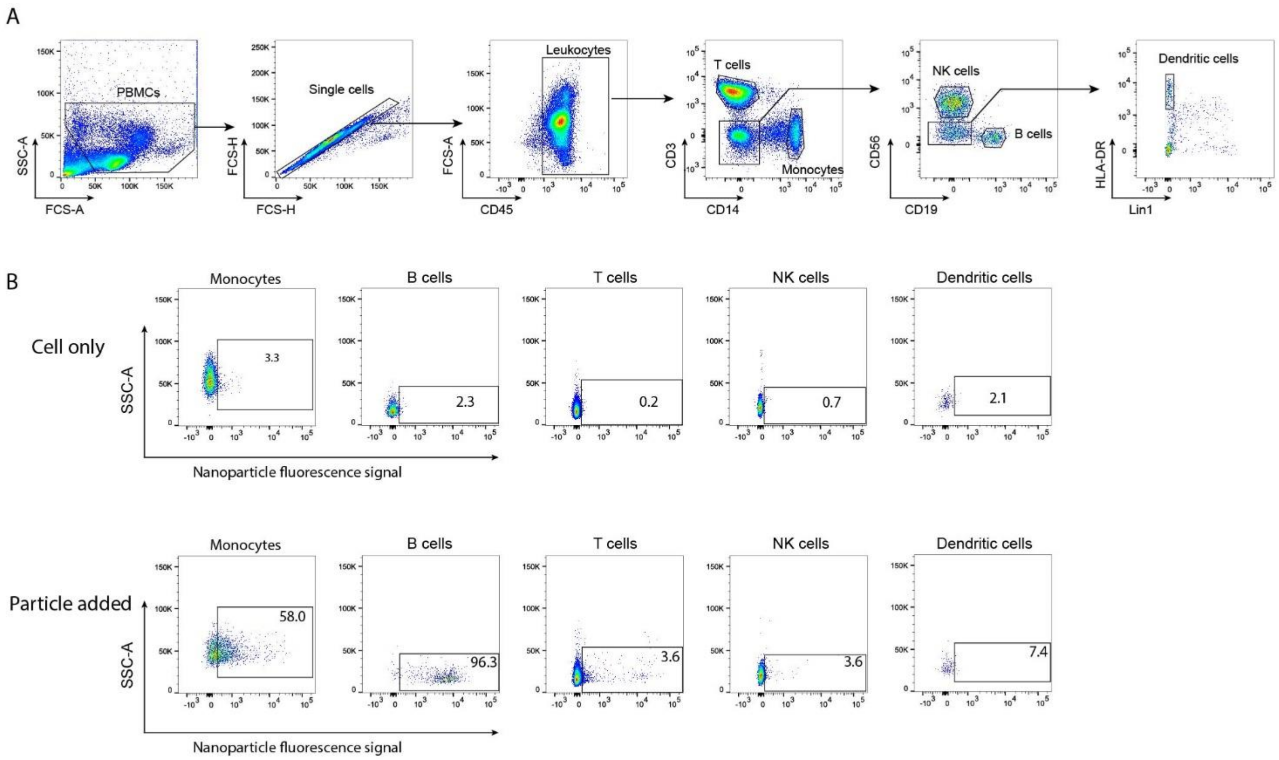


**Figure S8.** FlowJo gating strategy to analyze flow cytometry data. (a) Flow cytometry gating strategy used to identify different immune cell populations from human PBMCs. Forward and side scattering was used to locate white blood cells, followed by excluding doublets. The following cell types were identified based on the expression of surface markers: CD45+ leukocytes, CD14+ monocytes, CD3+ T cells, CD19+ B cells, CD56+ natural killer (NK) cells, and Lin-HLA-DR+ dendritic cells. (b) The percentage of each cell type positive for the fluorescence-labeled lipid nanoparticles was then measured as the cell association (%). The representative gating strategy and cell association percentages for each cell type is shown.

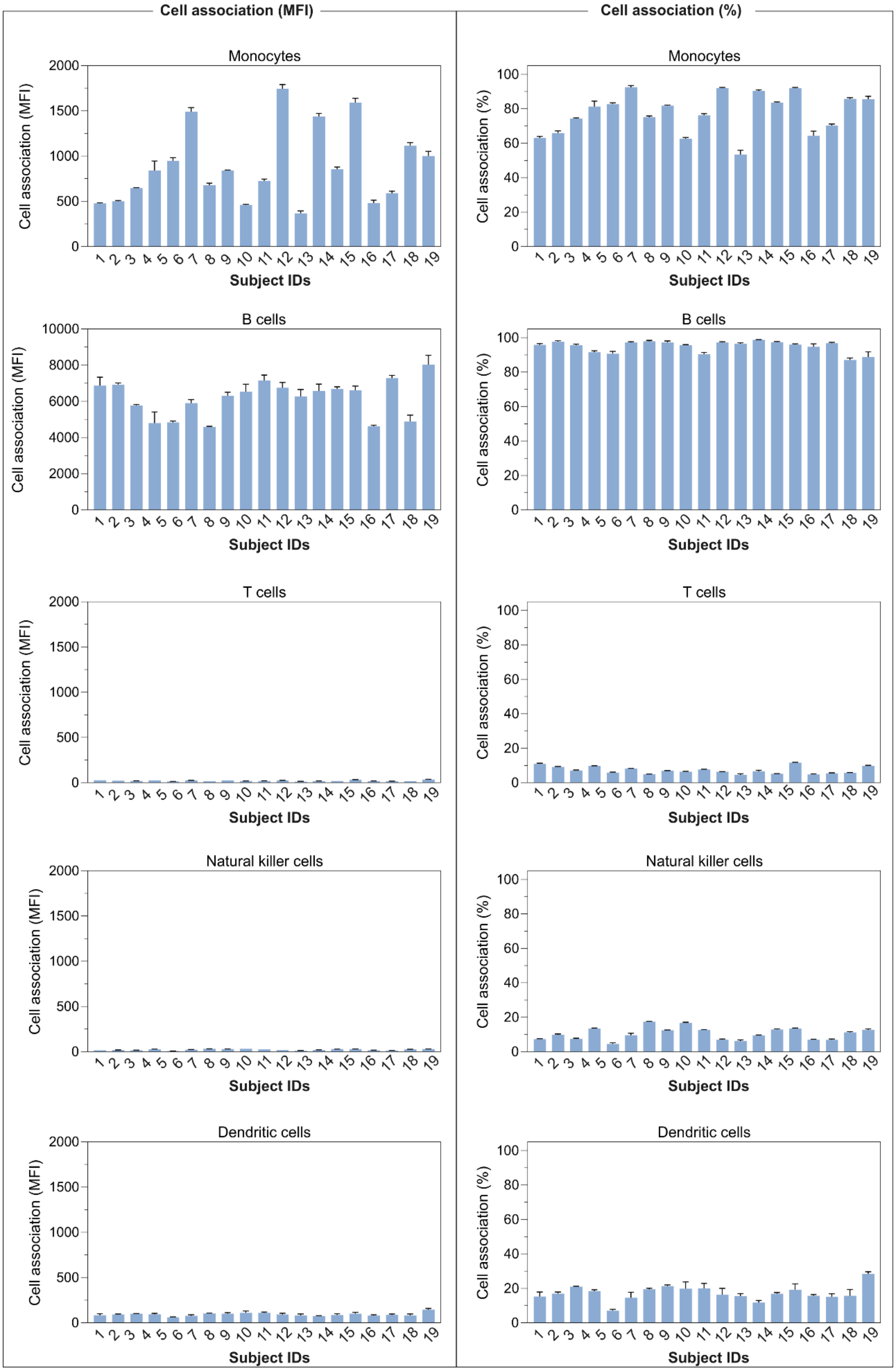

Cell association (MFI)
Cell association (%)
Monocytes
B cells
T cells
Natural killer cells
Dendritic cells
Cell association (MFI)
Cell association (%)
Subject IDs

**Figure S9.** Impact of the person-specific PBMCs on lipid nanoparticle−immune cell interactions. Lipid nanoparticles were pre-incubated with a human plasma (collected from 1 donor pre-vaccination) and then incubated with PBMCs from the 19 donors (collected pre-vaccination) of the Moderna bivalent vaccinee cohort (Table S1) in serum-free media for 1 h at 37 °C, followed by phenotyping cells with antibody cocktails and analysis by flow cytometry. Cell association (%) refers to the proportion of each cell type with positive fluorescence, above background, stemming from DiD-labeled particles. Cell association (MFI) refers to the median red fluorescence index of each cell type. Data are shown as the mean ± SD of three independent experiments, with at least 150,000 leukocytes analyzed for each experimental condition studied.

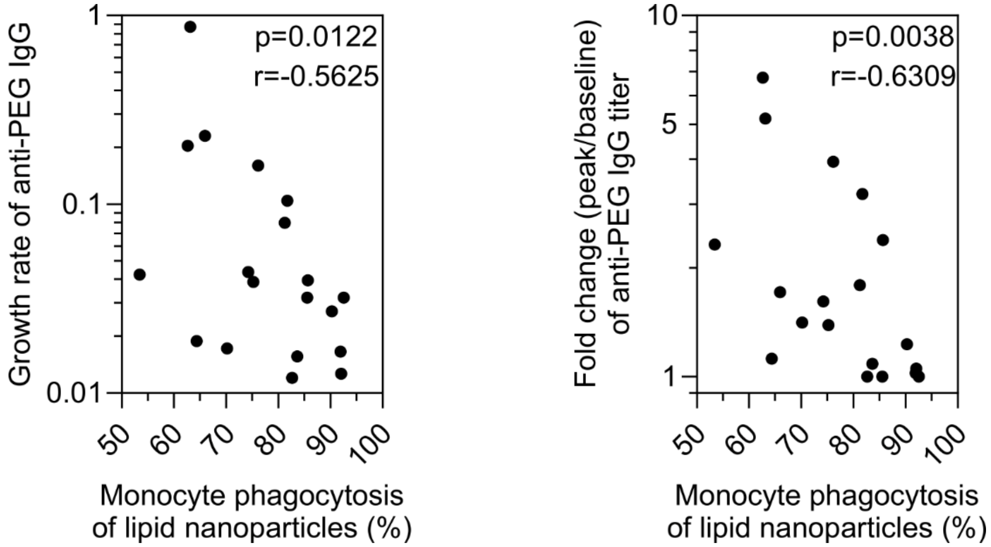


**Figure S10.** Significant negative correlation between monocyte phagocytosis of lipid nanoparticles (%) and anti-PEG IgG expansion (growth rate or fold change). Statistics were assessed by Spearman correlation analysis.

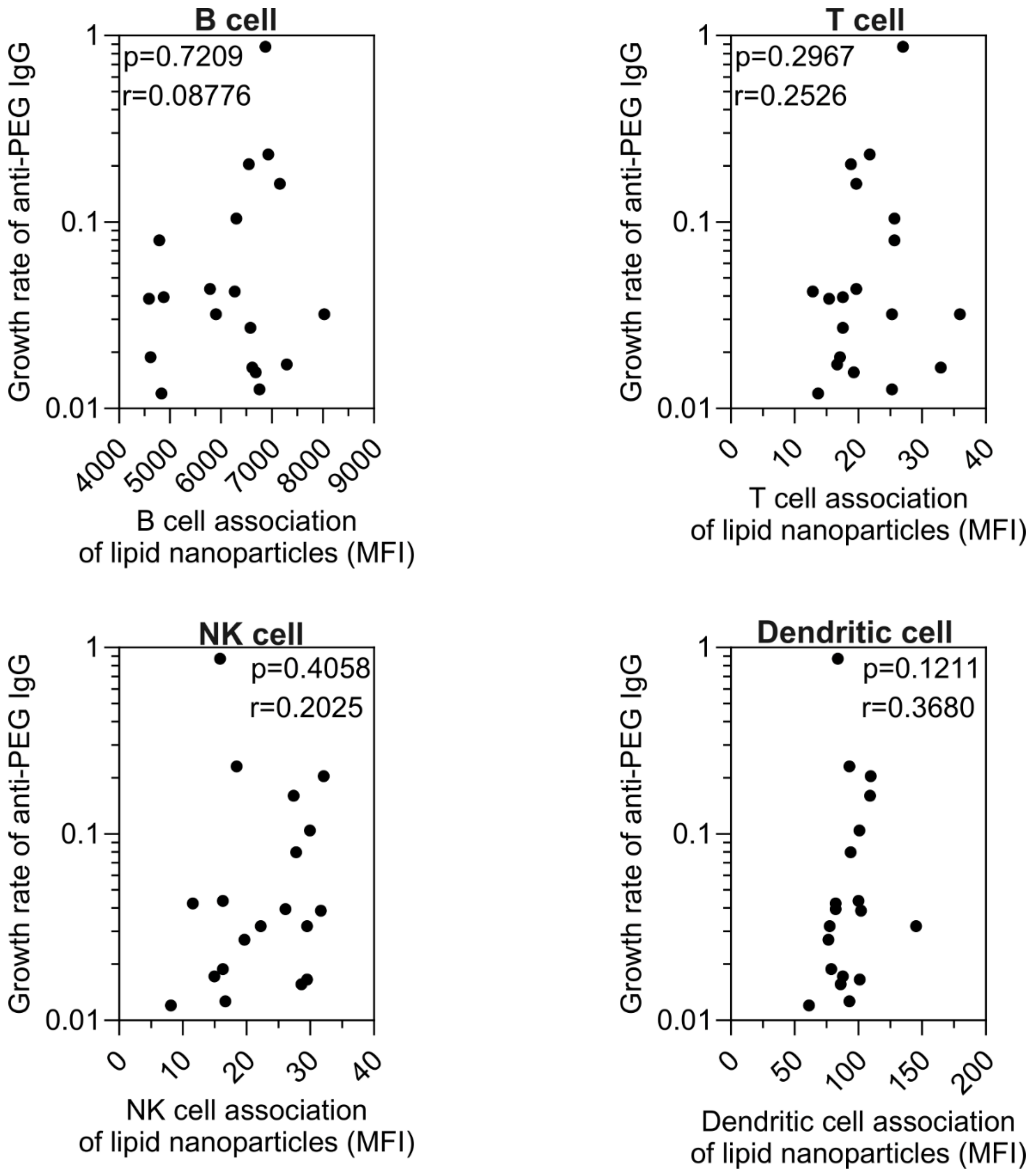


**Figure S11.** No correlation between B cell/T cell/NK cell/Dendritic cell association (MFI) and anti-PEG IgG expansion. Statistics were assessed by Spearman correlation analysis.
